# Cortico-Cortical Paired Associative Stimulation Increases SMA-M1 Facilitation in Tremor-Dominant Parkinson’s Disease

**DOI:** 10.64898/2026.04.28.26351457

**Authors:** Jane Tan, Brittany K. Rurak, Rick C. Helmich, Julian P. Rodrigues, Brian D. Power, Peter Drummond, Hakuei Fujiyama, Ann-Maree Vallence

**Author notes:** **Declarations of interest**: R.C.H. received funds for consultancy from Neurocrine Biosciences. **Corresponding Author:** Jane Tan, *Email address*, *Postal address*: Murdoch University, 90 South Street, Murdoch, WA 6150, Australia.

## Abstract

**Objective:** Tremor is a common motor symptom of Parkinson’s disease (PD) that reduces quality of life. Cortico-cortical paired associative stimulation (ccPAS) is a transcranial magnetic stimulation protocol that induces connective associative plasticity between cortical regions. This study investigated the effects of ccPAS over supplementary motor area (SMA) and primary motor cortex (M1) on SMA-M1 facilitation and tremor severity in individuals with tremor-dominant PD.

**Methods:** Fourteen individuals with tremor-dominant PD (mean age 66.5 ± 6.3 years; 5 females) received real and sham ccPAS in separate sessions. SMA-M1 activity and tremor severity were measured before and after ccPAS. Participants were tested OFF dopaminergic medication.

**Results:** SMA-M1 facilitation increased significantly after real but not sham ccPAS. There was no significant change in tremor severity after either real or sham ccPAS. Tremor severity was not associated with the change in SMA-M1 activity following ccPAS or with baseline SMA-M1 activity.

**Conclusions:** ccPAS can increase SMA-M1 facilitation in individuals with tremor-dominant PD. A single session of SMA-M1 ccPAS was insufficient in modulating tremor severity.

**Significance:** This study provides preliminary evidence of significant increases in SMA-M1 facilitation from ccPAS in tremor-dominant PD OFF dopamine medication. Future research should explore ccPAS effects ON dopamine medication, and the effects of multi-session ccPAS.

## 1. Introduction

Parkinson’s disease (PD) is a neurodegenerative disorder characterised by slowed movements and rigidity and/or tremor (Bloem et al., 2021). Tremor, a repetitive and involuntary shaking that typically occurs in the limb(s), is one of the most common motor symptoms of PD (Dirkx & Bologna, 2022). Tremor interferes with activities of daily living such as eating, writing, typing, and dressing, which may reduce quality of life (Heusinkveld et al., 2018). PD pathology involves the degeneration of nigrostriatal dopamine cells in the basal ganglia, and dopaminergic medication, such as levodopa, is the mainstay treatment for PD-related motor symptoms (Bloem et al., 2021). However, tremor severity is not related to nigrostriatal dopamine depletion, and the effects of levodopa on tremor can be inconsistent (Pirker et al., 2023; Zach et al., 2020). Therefore, the development of alternative or adjunct treatments for tremor is crucial.

The supplementary motor area (SMA) connects to the internal segment of the globus pallidus (GPi) within the basal ganglia via the striatum, receives inputs from GPi via the ventral lateral nucleus of the thalamus (Akkal et al., 2007; Strick, 1985), and has direct cortical projections to the primary motor cortex (M1) (Dum & Strick, 2002; Luppino et al., 1993). Both M1 and SMA are implicated in resting tremor: the delivery of single-pulse transcranial magnetic stimulation (TMS) to M1 reduces resting tremor intensity (Helmich et al., 2021) and TMS to M1 and SMA resets resting tremor (Lu et al., 2015). Previous research using diffusion tensor imaging and retrograde tracing has found M1 and SMA to be structurally connected (Muakkassa & Strick, 1979; Schulz et al., 2015). Furthermore, the ability of SMA to facilitate M1 activity is associated with lower resting tremor severity in individuals with tremor-dominant PD (Rurak et al., 2024).

The modulatory effects of SMA on M1 can be measured using a dual-site TMS protocol which involves a suprathreshold intensity conditioning stimulus (CS) delivered to SMA at interstimulus intervals (ISI) of 6-8 ms before a test stimulus (TS) is delivered to M1 (Arai et al., 2012; Rurak et al., 2021a, 2024). A larger motor evoked potential (MEP) amplitude elicited by dual-site paired-pulse trials compared to single-pulse trials (TS delivered to M1 alone) indicates that SMA exerts a facilitatory effect on M1 (Arai et al., 2012; Rurak et al., 2021a). This facilitatory effect of SMA on M1 is likely due to the glutamatergic interactions between SMA and M1 (Shima & Tanji, 1998). A recent study using dual-site TMS found that SMA-M1 modulation tends to be inhibitory in individuals with PD, which is atypical compared with healthy younger and older adults (Rurak et al., 2024). Notably, greater SMA-M1 facilitation significantly correlated with lower tremor severity for individuals with PD ON levodopa medication (Rurak et al., 2024). Based on these findings, it is plausible that increasing SMA-M1 facilitation could reduce tremor severity.

Cortico-cortical paired associative stimulation (ccPAS) is a repeated dual-site TMS protocol that can induce connective associative plasticity between cortical regions (Rizzo et al., 2009). During ccPAS, two cortical regions are repeatedly stimulated at fixed and constant ISI based on the temporal properties of short-latency connections between the regions (Hernandez-Pavon et al., 2023). Depending on the temporal order and ISI of the pulses applied to the target regions, ccPAS can induce Hebbian spike-timing-dependent plasticity (STDP; Rizzo et al., 2009). During Hebbian STDP, long-term potentiation (LTP) typically occurs when action potentials at excitatory presynaptic neurons are repeatedly followed by postsynaptic action potentials; conversely, long-term depression (LTD) usually occurs when presynaptic activation occurs after postsynaptic activation (Bi & Poo, 1998; Magee & Johnston, 1997; Markram et al., 1997). ccPAS involving the repeated stimulation of left M1 before right M1 at ISIs of 8 ms (but not 1 ms or random ISIs) was found to reduce left-to-right M1 interhemispheric inhibition and increase right M1 corticospinal excitability, suggesting LTP-like effects (Rizzo et al., 2009). A recent systematic review also suggests that ccPAS has the potential to modulate corticospinal excitability and cortico-cortical activity, with behavioural outcomes such as improved manual dexterity and visuomotor performance (Hernandez-Pavon et al., 2023).

Notably, ccPAS with SMA stimulated at an ISI of 6 ms before M1 stimulation significantly increased corticospinal excitability in healthy individuals (Arai et al., 2011). The ISI was set so that excitatory synaptic output from SMA would arrive at the same time or shortly before the generation of action potentials in M1 from M1 stimulation (Arai et al., 2011). Indeed, corticospinal excitability *decreased* when SMA was stimulated 15 ms *after* M1 stimulation, and there were no significant changes in corticospinal excitability when SMA was stimulated 10 ms after or 3.2 ms before M1 stimulation (Arai et al., 2011). However, the effects of ccPAS to SMA and M1 have not yet been explored in individuals with PD.

The current study explored the effects of ccPAS on the modulatory effects of SMA on M1 in individuals with tremor-dominant PD. We hypothesised that ccPAS to SMA and M1 would increase SMA-M1 modulation and decrease resting tremor intensity. We also conducted an exploratory correlational analysis to examine whether changes in SMA-M1 modulation were associated with changes in tremor severity. We hypothesised that increases in SMA-M1 facilitation would be significantly associated with decreases in tremor intensity.

## 2. Methods

### 2.1. Participants

Fourteen individuals with PD (mean age 66.5 ± 6.3 years; 5 females) participated in the study. All participants had idiopathic PD, resting tremor in at least one upper limb, and were treated with levodopa. Participants were excluded if they had contraindications to TMS or potential cognitive impairments (<23 on the Montreal Cognitive Assessment as recommended by Carson et al., 2018; mean score 28.71 ± 1.27). Individuals with head tremor or upper limb dyskinesia were also excluded to avoid difficulties with TMS coil placement and upper limb tremor measurement. Participant characteristics are shown in Supplementary Material Table S1. This study was performed in line with the ethical standards of the 1964 Helsinki Declaration and its later amendments or comparable ethical standards. The study was approved by the Murdoch University Human Research Ethics Committee (2018/117). All participants gave written informed consent.

### 2.2. Experimental Protocol

Figure 1A shows the experimental protocol. Each participant attended two 2-hour experimental sessions, one involving real ccPAS and the other involving sham ccPAS. All participants were tested in the OFF-medication state to examine ccPAS effects independent of levodopa. All experimental sessions were conducted in the morning (range: 8.30-9.30 am) to ensure ≥ 12 hours overnight withdrawal from levodopa (range: 12.5-15 hours). The sessions were counterbalanced across participants and separated by at least one week, and at the same time of day to minimise confounds from circadian changes in cortisol levels on corticospinal excitability (Sale et al., 2008). Seven participants received real ccPAS in their first session, and seven other participants received sham ccPAS in their first session. During each session, resting tremor intensity and SMA-M1 modulation were measured before and after the delivery of ccPAS.

**Figure 1.**
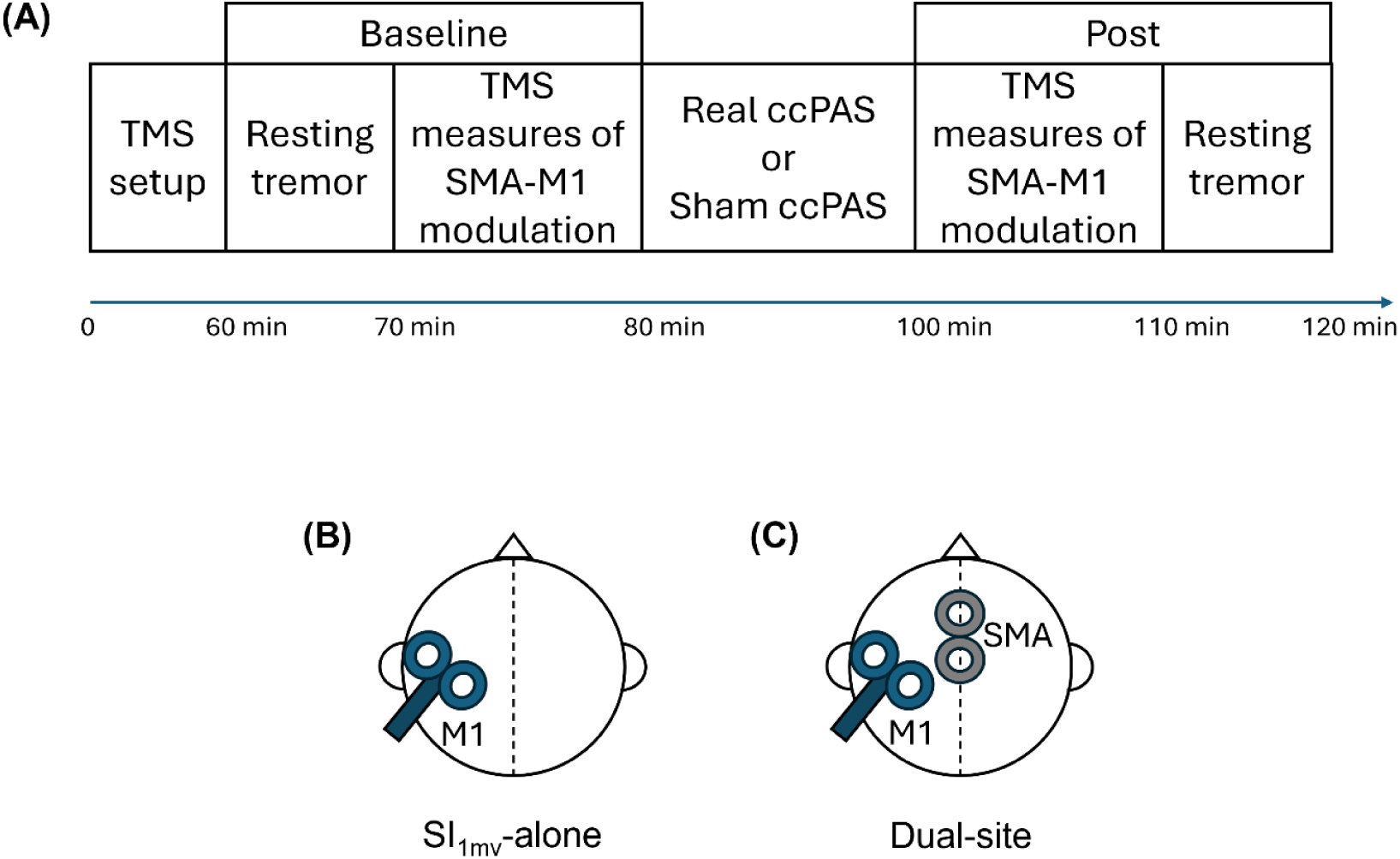
The Experimental Protocol. *Note*. (A) All participants attended two sessions, one involving real ccPAS and the other involving sham ccPAS. (B) During single-pulse trials, stimulation was applied only to M1 (SI_1mv_-alone). M1 stimulation coil was placed on the M1 FDI hotspot contralateral to the tremor-dominant arm. (C) During dual-site trials, a conditioning pulse was delivered to SMA at an ISI of 7 ms before the test pulse to M1. The SMA stimulation coil was placed on the midline, 4 cm anterior to the vertex, in a lateral orientation.

Resting tremor was also measured during a provocation task: participants performed a serial 7 subtraction task. Resting tremor is typically amplified by cognitive load (Dirkx et al., 2020), and we were interested in examining whether ccPAS differentially affected resting tremor with and without provocation. The analysis and results for resting tremor during the provocation task are reported in Supplementary Material Section S2.

### 2.3. TMS

EMG activity was recorded from the relaxed first dorsal interosseous (FDI), extensor carpi radialis (ECR), and flexor carpi radialis (FCR) muscles of the tremor-dominant upper limb, using Ag-AgCI surface electrodes in a belly-tendon montage. The EMG signal was amplified (x1000; CED 1902), bandpass filtered at 2-1000Hz, notch-filtered at 50Hz, and digitised at 5kHz (CED 1401). Dual-site TMS was delivered to SMA and M1 using two 50-mm figure-of-eight coils each connected to a Magstim 200^2^ stimulator (Magstim Co., UK).

The M1 stimulation coil was placed on the M1 FDI hotspot contralateral to the tremor-dominant arm (Figure 1B; n = 6 left arm). The coil was held tangentially to the scalp with the handle positioned backwards, away from the midline by ∼45◦ to induce a posterior-anterior current. M1 stimulation intensity was set as the intensity (as a percentage of maximum stimulation output; %MSO) that elicited MEPs of ∼1 mV peak-to-peak amplitude (SI_1mV_) in the resting FDI.

The SMA stimulation coil was placed on the midline, 4 cm anterior to the vertex, in a lateral orientation (Figure 1C) (Arai et al., 2011, 2012; P. E. Green et al., 2018; Rurak et al., 2021b, 2021a, 2024). SMA stimulation intensity was set at 140% of the active motor threshold (AMT). The AMT was defined as the minimum intensity (%MSO) that elicited MEPs in FDI with amplitudes ≥0.2 mV in 5/10 consecutive trials during an isometric contraction of 10% maximum voluntary contraction (Jitkritsadakul et al., 2016).

To avoid coil overheating, two separate sets of coils were used to measure SMA-M1 modulation and deliver ccPAS. Stimulation intensities were determined for both sets of coils in both experimental sessions for each participant.

During TMS measures of SMA-M1 modulation, coil position was monitored in real-time using neuronavigation software (Brainsight TMS, Rogue Research, Canada), with the Montreal Neurological Institute International Consortium for Brain Mapping (MNI ICBM) 152 average brain template. The nasion, and left and right preauricular points were used as landmarks for patient-to-image registration, with scaling landmarks at ∼1 cm steps along anterior-lateral and anterior-posterior. Registration verification was confirmed with a distance of 3 mm or less.

Neuronavigation was not used during ccPAS as the tops of the ccPAS coils were covered by pre-chilled heat sinks (Belyk et al., 2019) to avoid overheating, thus preventing neuronavigation sensors from being attached.

#### 2.3.1. SMA-M1 Ratio

SMA-M1 modulation was measured with single-pulse and dual-site trials. During single-pulse trials, stimulation was applied to M1 only (SI_1mv_-alone). During dual-site trials, a conditioning pulse was delivered to SMA (140% AMT) at an ISI of 7 ms (Rurak et al., 2021a) before the test pulse to M1. SI_1mv_-alone and dual-site trials were pseudo-randomised with an inter-trial interval of 5s (±10%). Two experimental blocks involving 30 trials each (15 SI_1mv_-alone and 15 dual-site) were delivered. Each block lasted ∼3 mins.

A custom Signal script was used to trigger TMS pulses if EMG activity was within individualised EMG thresholds for a period of 50 ms (horizontal dotted lines in Figure 2A). The individualised EMG thresholds were determined from baseline (pre-ccPAS) measurements of resting tremor, using EMG activity from the muscle showing the clearest tremor bursts (tremor-dominant muscle). If EMG activity was not below thresholds after 50 ms, TMS pulses were delivered after ∼ 5 s (±2%) (Figure 2B) to limit the length of the experimental sessions. These trials were excluded from analyses. There were no significant differences in the mean number of trials excluded between real (*M* = 2.00, *SD* = 3.09) and sham (*M* = 4.35, *SD* = 6.64) sessions (*p* = .138).

**Figure 2.**
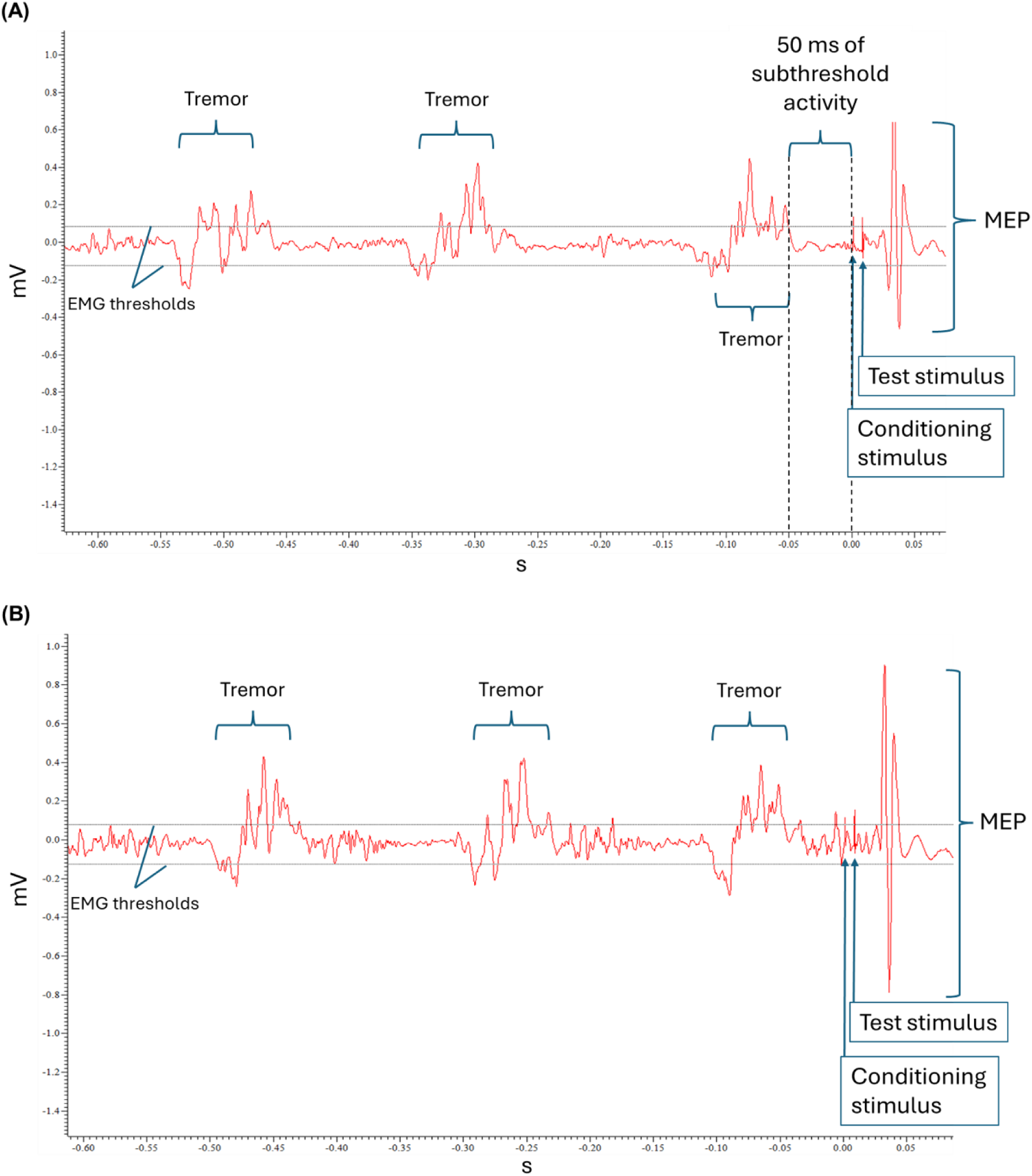
Example EMG Traces for TMS trials. *Note*. Example EMG traces for TMS trials. SI_1mV_-alone and dual-pulse trials were delivered: (A) after 50ms of EMG activity within EMG thresholds (horizontal dotted lines) determined for each participant, or (B) if a period of 50ms passed without subthreshold EMG activity – these trials were removed from analysis. Thresholds were established for each participant from baseline tremor activity, before TMS measurement.

Participants were instructed to be silent and alert with their eyes opened and not to suppress tremor activity during TMS (Groppa et al., 2012; Pellegrini et al., 2020).

SMA-M1 modulation was measured by the ratio of the mean dual-site MEP amplitude to the mean SI_1mv_-alone MEP amplitude. Ratios larger than 1 denote a facilitatory effect of SMA stimulation on M1, and ratios smaller than 1 denote an inhibitory effect of SMA stimulation on M1.

#### 2.3.2. ccPAS

ccPAS involved dual-site trials where a conditioning pulse was delivered to SMA (140% AMT) at an ISI of 7 ms before a test pulse to M1 (SI_1mV_). An inter-trial interval of 5 s (0.2 Hz) was used. The stimulation parameters were based on the M1-SMA ccPAS protocol in Arai et al. (2011). We utilized a 7-ms ISI as a previous study found moderate test-retest reliability when the ISI was 7 ms, and poor reliability when ISIs were 6 ms and 8 ms (Rurak et al., 2021a). To prevent overheating of the coils, ccPAS was administered in three blocks of 50 trials each, separated by 2-minute intervals (Arai et al., 2011). During sham ccPAS, only M1 was effectively stimulated: the SMA stimulation coil was held perpendicular to the scalp to avoid direct stimulation of the SMA site (Boylan et al., 2001). Condensation-free pre-chilled heat sinks (Belyk et al., 2019) were also used to prevent the TMS coils from overheating during ccPAS application.

### 2.4. Resting Tremor

Surface electromyographic (EMG) activity was recorded from FDI, ECR, and FCR muscles in the tremor-dominant limb, using Ag-AgCI surface electrodes in a belly-tendon montage. During tremor measurement, participants kept their tremor-dominant forearm relaxed on the chair armrest with their hand unsupported and not touching anything. Tremor measurement was conducted in three 1-minute blocks. The EMG signal was amplified (x1000; CED 1902), bandpass filtered (2-500Hz), notch-filtered (50Hz), and digitised at 5kHz (CED 1401). After data acquisition, the EMG signal was down-sampled (1000Hz), rectified, and bandpass filtered (second-order Butterworth filter; 2-11Hz).

### 2.5. Data analysis

All analyses were performed using R (version 4.3.2) and RStudio (version 2024.12.0+467), using packages *tidyverse*, *readxl*, *ggpubr*, *rcompanion*, *emmeans*, *skimr*, *lme4*, *lmerTest*, *car*, *rstatix*, *glmm*, and *irr* (Bates et al., 2015, 2025; Fox et al., 2024; Fox & Weisberg, 2019; Gamer et al., 2019; Kassambara, 2021, 2023; Knudson, 2024; Kuznetsova et al., 2017; Lenth, 2023; Mangiafico, 2024; Waring et al., 2022; Wickham, 2023; Wickham & Bryan, 2023).

As the trial-level data for EMG activity in the 50 ms immediately preceding TMS (pre-TMS RMS; see next section 3.1.1) and MEP amplitudes of SI_1mv_-alone trials had non-negative and positively-skewed distributions, they were analysed with generalised linear mixed models (GLMMs), with a gamma distribution and log link function applied (Puri & Hinder, 2022).

SMA-M1 ratios and resting tremor power were analysed using linear mixed models (LMMs). All GLMMs and LMMs were analysed with the maximal random effect structure (by-participant random intercepts, by-participant random slopes for all fixed effects, and correlations among random effects) as allowed by the data and justified by the design used for every model (Barr et al., 2013; Singmann & Kellen, 2019). Null hypothesis significance testing for main and interaction effects was conducted using Wald Chi-Squared tests for GLMMs and *F*-tests for LMMs. Significant effects were investigated with Bonferroni-corrected contrasts. Effect sizes for post-hoc contrasts were measured with Cohen’s *d*, with *d* ≈ 0.2 for a small effect, *d* ≈ 0.5 for a medium effect, and *d* ≈ 0.8 for a large effect (Cohen, 1988). Statistical significance was accepted at an alpha value of .05.

#### 2.5.1. Neurophysiological Measures

EMG activity was recorded from FDI, ECR, and FCR during TMS delivery. However, as the stimulation parameters were optimised for the FDI, only EMG activity from the FDI was analysed.

Paired-samples *t* tests were conducted to compare the stimulation intensities between real and sham ccPAS sessions.

To ensure that the effects of ccPAS on SMA-M1 modulation were not confounded by background EMG activity from tremor, EMG activity in the 50 ms immediately preceding TMS (pre-TMS RMS) was analysed with a GLMM, with fixed effects of STIMULATION (real, sham), TIME (pre, post ccPAS), and TRIALTYPE (SI_1mv_-alone, dual site).

The MEP amplitudes of SI_1mv_-alone trials were analysed with a GLMM, with fixed factors of STIMULATION (real, sham) and TIME (pre, post).

#### 2.5.2. SMA-M1 Ratio

SMA-M1 ratios were analysed for 10 participants; data from four participants could not be analysed due to a technical error. SMA-M1 ratios were analysed using a LMM with fixed factors of STIMULATION (real, sham) and TIME (pre, post). As the stimulation parameters were optimised for the FDI, only EMG activity from the FDI was analysed (see Supplementary Material Section S3 for ECR and FCR analyses).

#### 2.5.3. Resting Tremor

Welch’s power spectral density analysis was performed on each resting tremor block, using fast Fourier transform with a 0.04 Hz resolution. The spectral estimates were then averaged across the three blocks to obtain the average power of the tremor peak frequency for each individual. The resultant tremor power was log-transformed and analysed with a LMM with fixed factors of STIMULATION (real, sham), TIME (pre, post), and MUSCLE (FDI, ECR, FCR).

#### 2.5.4. Correlation Between SMA-M1 Ratio and Tremor Power

The effects of non-invasive brain stimulation protocols can be subject to inter-individual variability (López-Alonso et al., 2014). To account for potential inter-individual variability, we conducted an exploratory correlational analysis to examine whether changes in SMA-M1 modulation were associated with changes in tremor severity: the correlation between pre-post changes in SMA-M1 ratio and pre-post changes in tremor power was analysed using Spearman’s rank correlation coefficient (*ρ*) separately for real and sham ccPAS sessions. The correlation between baseline SMA-M1 ratio and baseline tremor power was also analysed using Spearman’s *ρ* separately for real and sham ccPAS sessions.

## 3. Results

### 3.1. Neurophysiological Measures

#### 3.1.1. Stimulation Intensities

There were no significant differences in stimulation intensities between real and sham ccPAS sessions for M1 (coil 1: *t*(13) = –0.71, *p* = .493; coil 2: *t*(13) = –1.09, *p* = .297) and SMA (coil 1: *t*(13) = 0.74, *p* = .474; coil 2: *t*(13) = 0.57, *p* = .578).

#### 3.1.2. Pre-TMS RMS

Figure 3 shows the trial-level pre-TMS RMS for each trial type, stimulation condition, and time point. The GLMM analysis of pre-TMS RMS found a significant TIME X STIM interaction (*χ*^2^(1, *N* = 10) = 5.19, *p* = .023). Post-hoc analyses showed a significant increase in pre-TMS RMS across both SI_1mV_-alone and dual-site trials after real ccPAS (*z* = –2.36, *p* = .018, *d* = –0.62), and no significant changes after sham ccPAS (*z* = –1.06, *p* = .290, *d* = – 0.26). There were no significant differences between real ccPAS and sham ccPAS for baseline pre-TMS RMS (*z* = –0.72, *p* = .472, *d* = –0.13) and pre-TMS RMS following ccPAS (*z* = 1.71, *p* = .088, *d* = 0.24). There were no other significant main effects or interactions (*χ^2^*s ≤ 3.34, *p*s ≥ .068).

**Figure 3.**
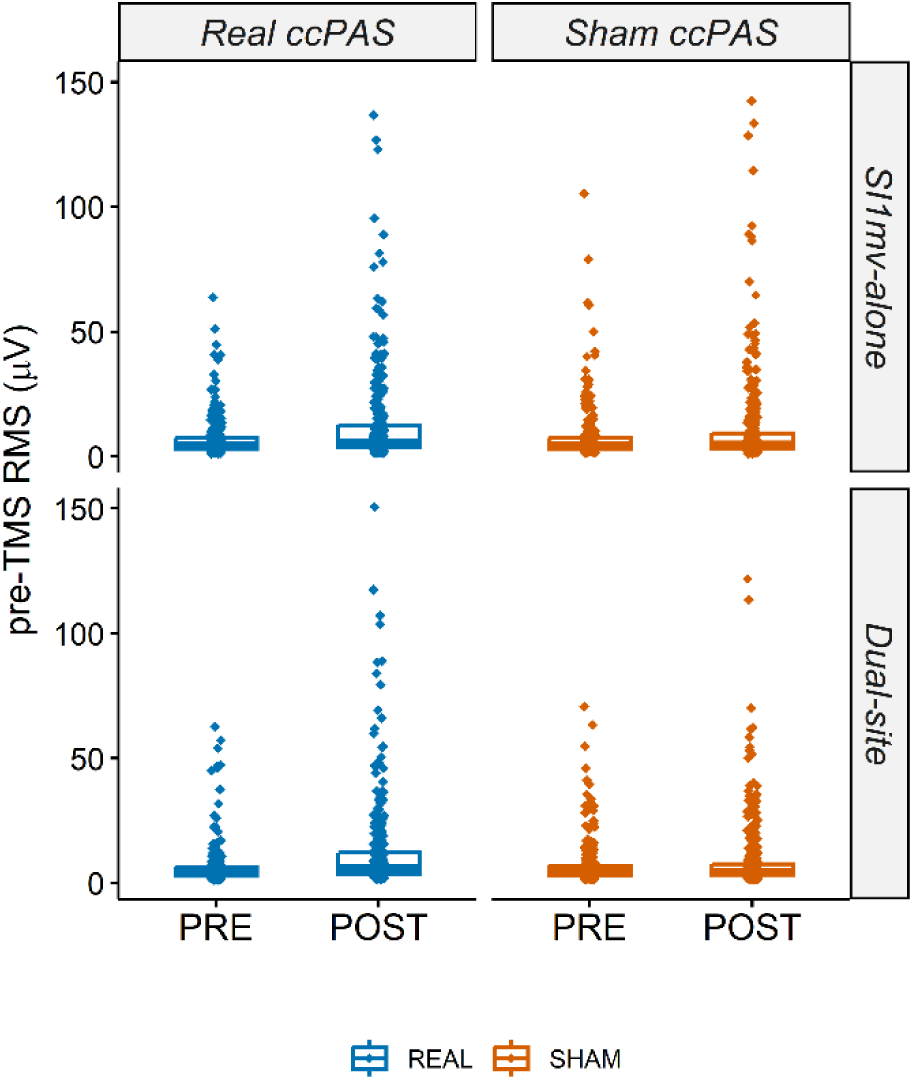
pre-TMS RMS. *Note*. Trial-level pre-TMS RMS for SI_1mV_-alone and dual-site trials before and after real and sham ccPAS.

To further investigate the significant increase in pre-TMS RMS during real ccPAS sessions, we examined the association between pre-TMS RMS and MEP amplitudes for each trialtype (SI_1mV_-alone and dual-site trials) and stimulation condition (real and sham ccPAS) (see Supplementary Material Section S4). During real ccPAS sessions, larger pre-TMS RMS was significantly associated with larger MEP amplitudes for SI_1mV_-alone (*ρ* = 0.20, *p* < .001) and dual-site trials (*ρ* = 0.19, *p* = .001.) at baseline but not after real ccPAS (*ρ* ≤ 0.058, *p*s ≥.329). Pre-TMS RMS was not associated with MEP amplitudes before or after sham ccPAS for SI_1mV_-alone and dual-site trials (*ρ* ≤ 0.11, *p*s ≥ .084).

#### 3.1.3. SI_1mV_-Alone MEP

Figure 4 shows the mean MEP amplitudes from SI_1mV_-alone trials for each stimulation condition at each timepoint. The GLMM analysis of SI_1mV_-alone MEP amplitudes found a significant main effect of TIME (*χ*^2^(1, *N* = 10) = 5.85, *p* = .016) across both stimulation conditions, with significant pre-post increases in MEP amplitudes for real and sham ccPAS conditions. There were no other significant main effects or interactions (*χ^2^*s ≤ 1.13, *p*s ≥.288).

**Figure 4.**
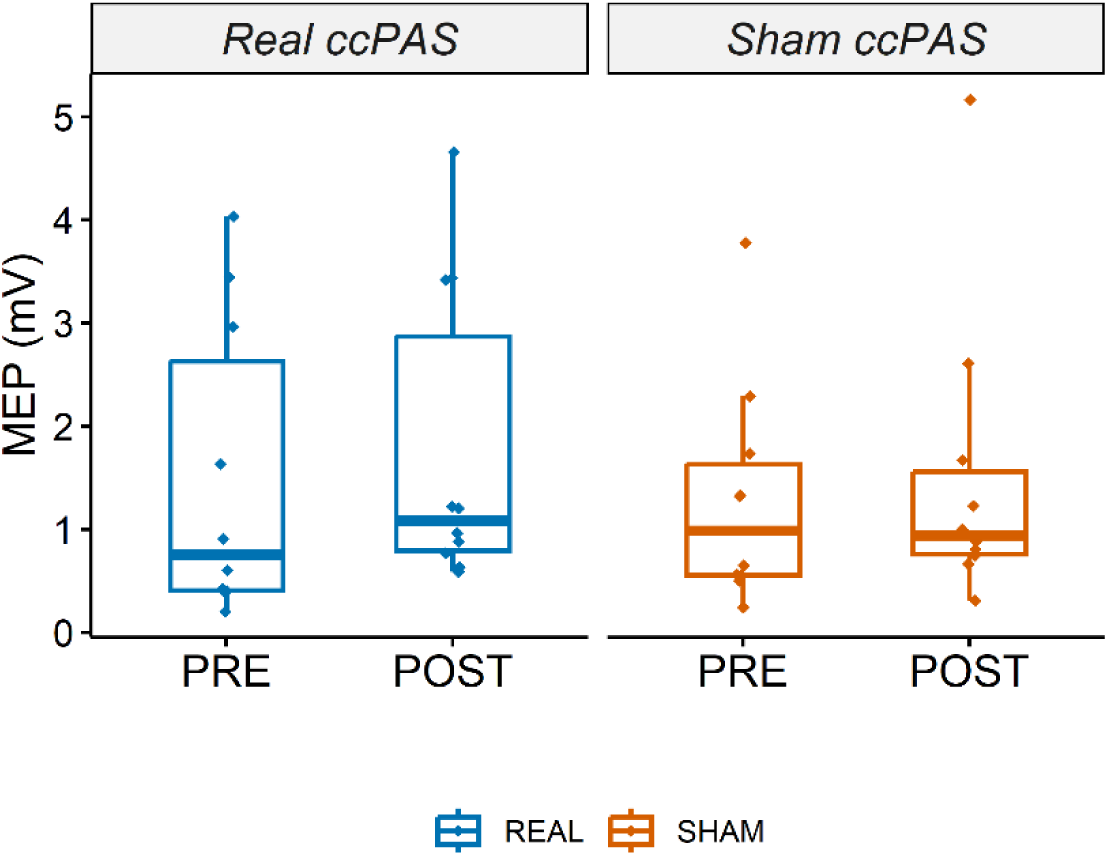
SI1mV-alone MEP. *Note*. Mean MEP amplitudes on SI_1mV_-alone trials before and after real and sham ccPAS

### 3.2. SMA-M1 Ratios

Figure 5 shows the SMA-M1 ratios for each stimulation condition at each timepoint. The LMM analysis of SMA-M1 ratios found a significant STIMULATION X TIME interaction (*F*(1, 11.11) = 6.16, *p* = .030). Post-hoc analyses showed a significant pre-post increase in SMA-M1 ratios for real ccPAS (*t* = –3.31, *p* = .004, *d* = –2.20), and no significant pre-post change in SMA-M1 ratios for sham ccPAS (*t* = –0.82, *p* = .425, *d* = –0.54). Baseline SMA-M1 ratios were lower for real ccPAS compared with sham ccPAS (*t* = –2.94, *p* = .009, *d* = –1.86). Post-ccPAS SMA-M1 ratios were not significantly different between real and sham ccPAS sessions (*t* = –0.32, *p* = .754, *d* = –0.20). The main effect of TIME was significant (*F*(1, 11.11) = 5.69, *p* = .036), with larger SMA-M1 ratios at post-ccPAS relative to baseline. The main effect of STIMULATION was not significant (*F*(1, 11.11) = 3.67, *p* = .082).

**Figure 5.**
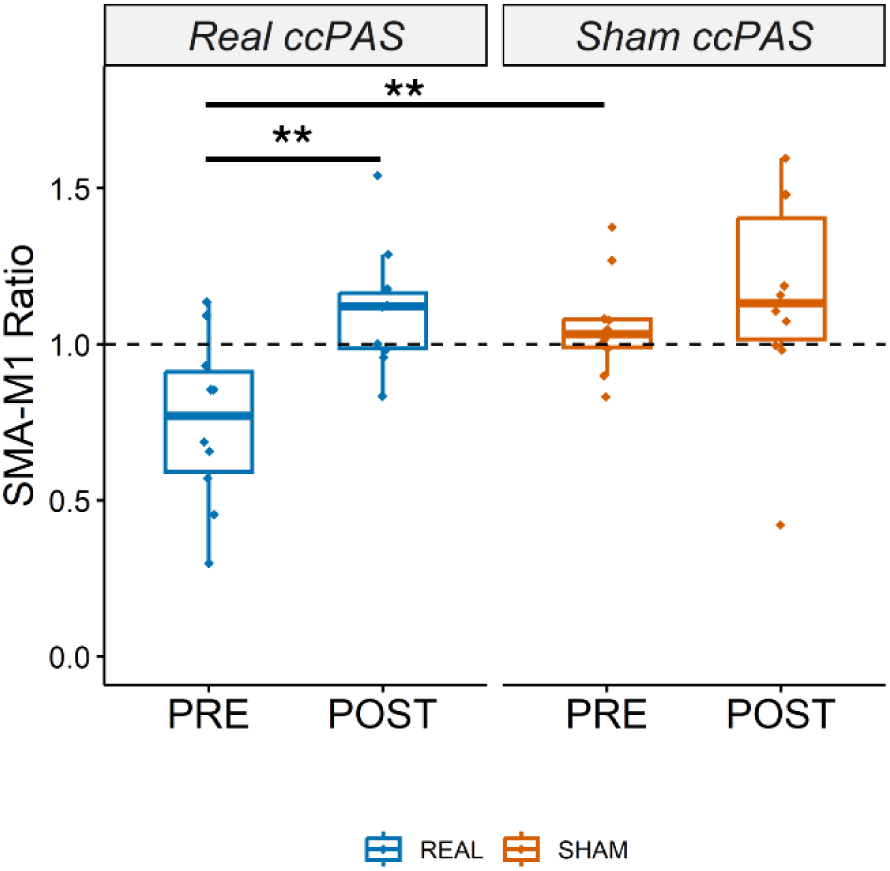
SMA-M1 ratios. *Note*. SMA-M1 ratios increased significantly after real ccPAS, and not sham ccPAS. \*\**p* <.01.

Given the significant difference in baseline SMA-M1 ratios between real and sham ccPAS sessions, we performed two exploratory analyses. First, we examined the association between baseline SMA-M1 ratios in the two sessions: there was no significant correlation between baseline SMA-M1 ratios in the real ccPAS condition and baseline SMA-M1 ratios in the sham ccPAS condition (Figure 6). Second, we examined the association between baseline SMA-M1 ratios and change in SMA-M1 ratio post ccPAS and found that smaller baseline SMA-M1 ratios were associated with larger pre-post increases in SMA-M1 ratios for real ccPAS but not sham ccPAS (see Figure 7).

**Figure 6.**
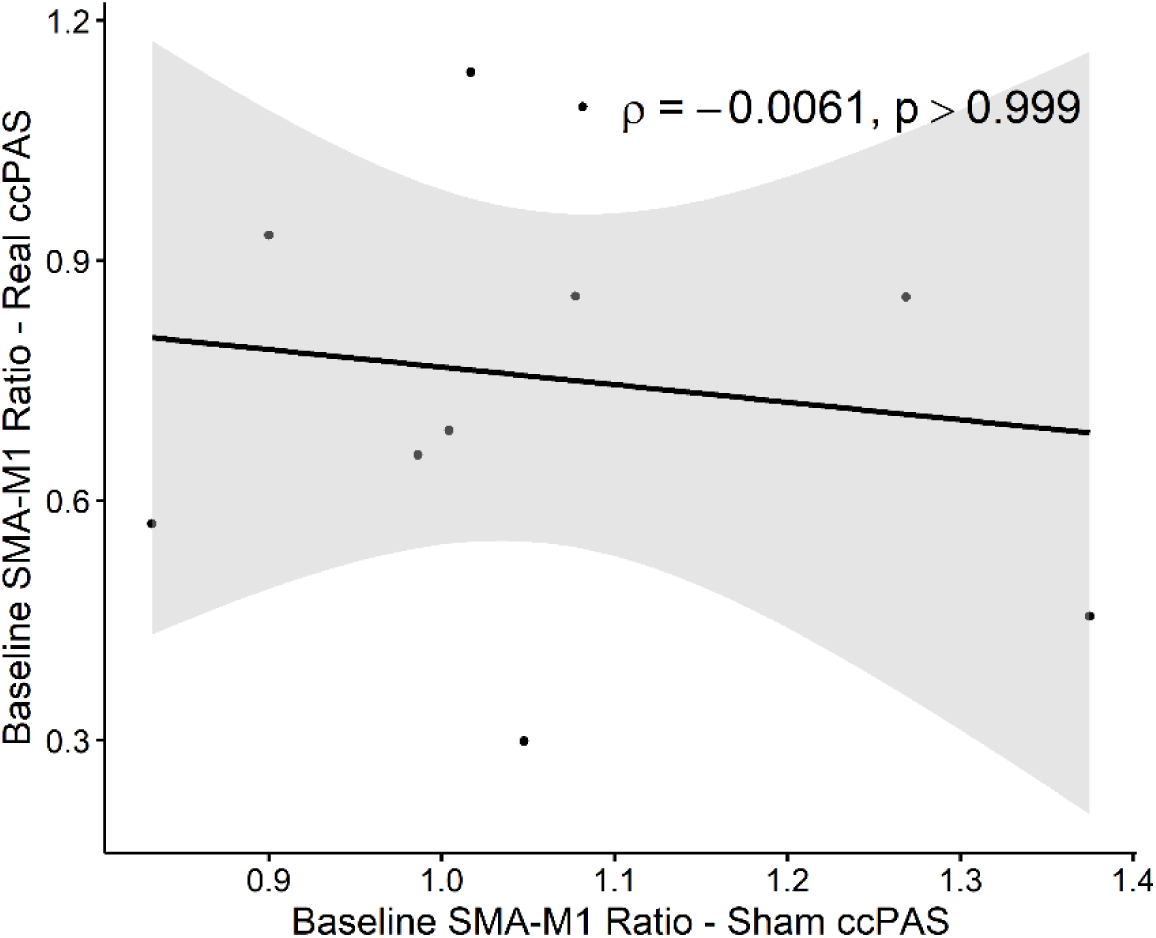
Scatterplot for Correlation between Baseline SMA-M1 ratios During Real and Sham ccPAS Sessions. *Note*. There was no significant correlation in baseline SMA-M1 ratios between real and sham ccPAS sessions.

**Figure 7.**
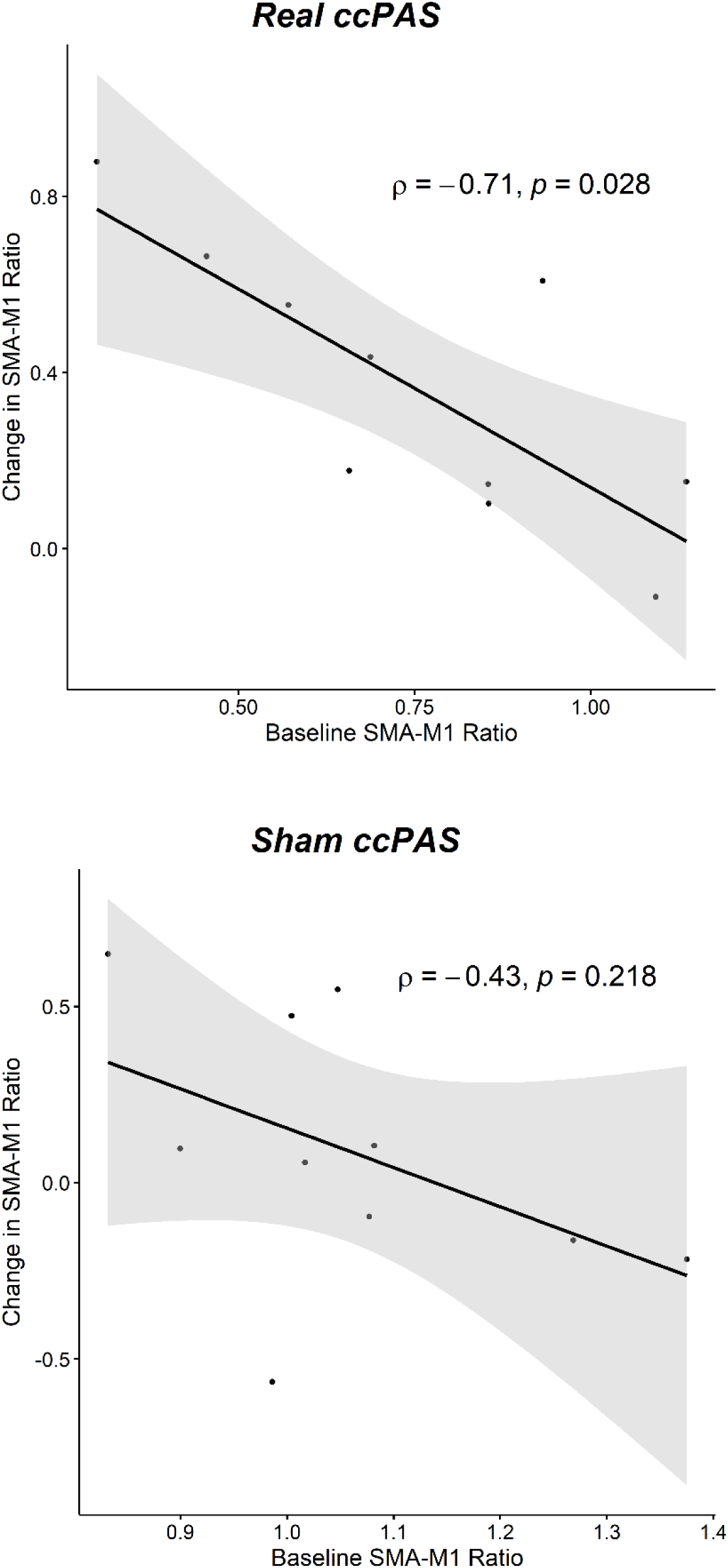
Scatterplots for Correlation between Baseline SMA-M1 ratios and Pre-Post Changes in SMA-M1 ratios. *Note*. During real ccPAS sessions, smaller baseline SMA-M1 ratios were significantly associated with larger increases in SMA-M1 ratios following ccPAS (top panel). Baseline SMA-M1 ratios were not significantly associated with changes in SMA-M1 ratios during sham ccPAS sessions (bottom panel).

### 3.3. Resting Tremor

Figure 8 shows the resting tremor power for each stimulation condition at each timepoint. The LMM analysis of resting tremor power found no significant interaction effects or main effects for STIMULATION, TIME, or MUSCLE (*F*s ≤ 2.33, *p*s ≥ .101). This was supported by a supplementary Bayesian LMM analysis of resting tremor power, which found no clear evidence for or against a practically meaningful effect of ccPAS on tremor power across stimulation types and muscles (Supplementary Material Section S6).

**Figure 8.**
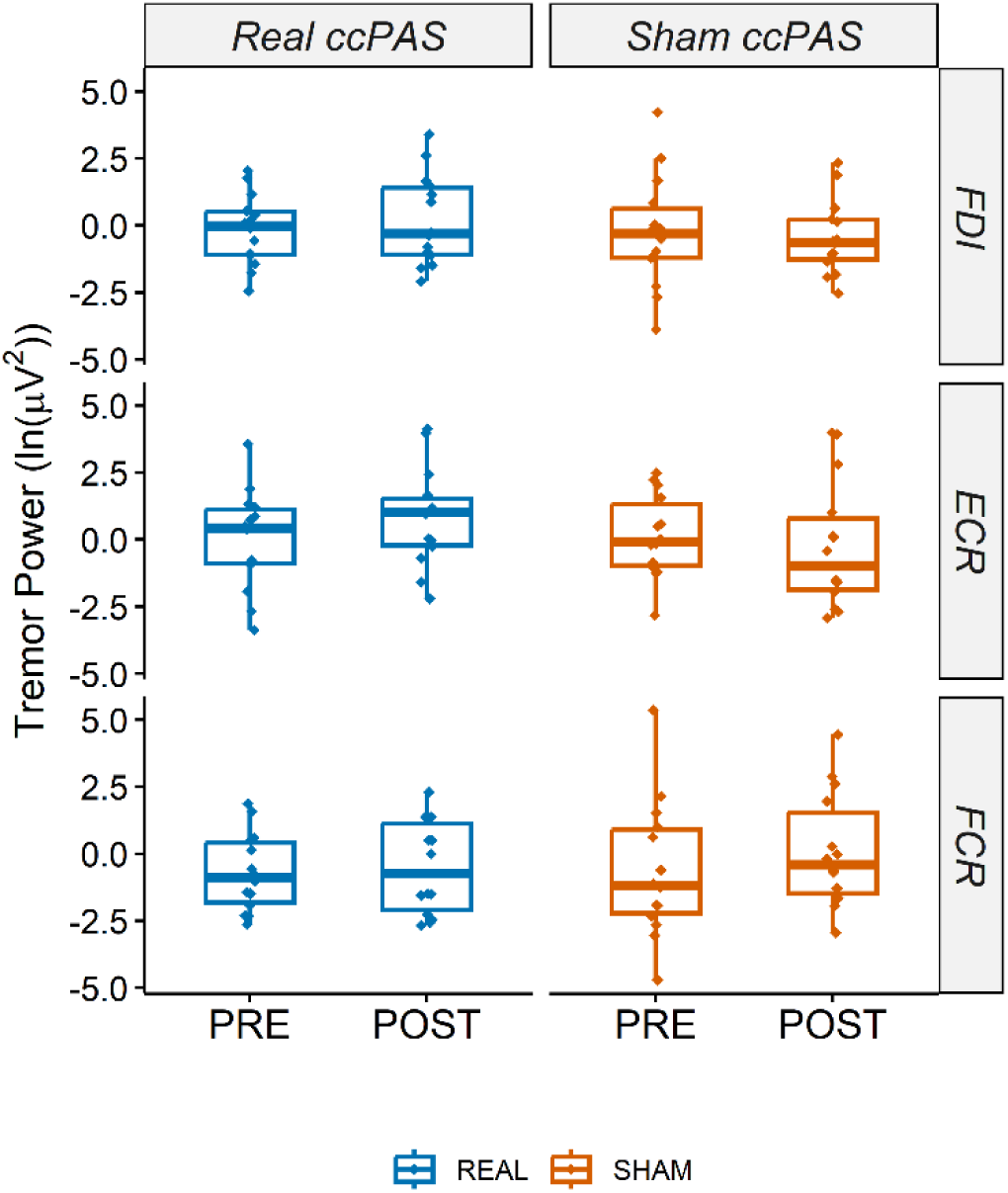
Resting Tremor Power. *Note*. There were no significant changes in resting tremor power from either real or sham ccPAS

A separate LMM analysis was also performed on resting tremor power for the 10 participants whose SMA-M1 ratios were analysed (see Supplementary Material Section S5). This found a significant main effect of TIME (*F*(1, 111.11) = 4.72, *p* = .032), with a pre-post increase in resting tremor power across both real and sham ccPAS sessions. There were no other significant interaction effects or main effects (*F*s ≤ 2.93, *p*s ≥ .056).

### 3.4. Correlation Between SMA-M1 Ratios and Tremor Severity

The exploratory correlational analysis found no significant correlation between pre-post changes in SMA-M1 ratios and pre-post changes in tremor power for either real or sham ccPAS sessions (*ρ* ≤ 0.13, *p*s ≥ .733; Supplementary Material Section S7 Figure S9). The correlation between baseline SMA-M1 ratios and baseline tremor power was also not significant for either real or sham ccPAS sessions (*ρ* ≤ –0.39, *p*s ≥ .263; Supplementary Material Section S7 Figure S10).

## 4. Discussion

This study investigated the effects of SMA-M1 ccPAS on SMA-M1 modulation and resting tremor severity in individuals with tremor-dominant PD. We found significant increases in SMA-M1 ratios after real ccPAS but not sham ccPAS. Contrary to our hypotheses, tremor severity did not change after ccPAS, and pre-post changes in SMA-M1 ratios were not associated with pre-post changes in tremor power.

### 4.1. ccPAS Increased SMA-M1 Facilitation

To our knowledge, this is the first study to provide preliminary evidence that ccPAS can effectively strengthen SMA-M1 facilitation in individuals with tremor-dominant PD. These results extend on previous work that showed facilitatory effects of SMA-M1 ccPAS in healthy younger adults (Arai et al., 2011) to suggest that ccPAS can induce STDP in individuals with PD, leading to a significant increase in SMA-M1 facilitation.

As SI_1mV_-alone MEP amplitudes increased across both real and sham ccPAS, the stimulation-specific increase in SMA-M1 facilitation was independent of an overall increase in corticospinal excitability. Since only M1 was effectively stimulated during sham ccPAS, its effects should be equivalent to that of a low-frequency (0.2 Hz) repetitive TMS (rTMS) protocol. Most studies have applied low-frequency rTMS at 1 Hz, which decreases corticospinal excitability (Fitzgerald et al., 2006). The effects of 0.2 Hz rTMS are less clear, but the current findings suggest that it might have a facilitatory effect on corticospinal excitability.

In the current study, there was a significant difference in baseline SMA-M1 ratios between sessions, specifically, smaller ratios before real ccPAS compared with sham ccPAS. At baseline, the experimental conditions for real ccPAS and sham ccPAS sessions were identical with no significant differences in stimulation intensities, therefore, the significant difference in baseline ratios between sessions is indicative of high intra-individual variability in SMA-M1 modulation. Previous research has shown greater temporal (moment-to-moment) variability in sensorimotor blood oxygen level-dependent activity for PD compared to healthy controls (Zhu et al., 2019), which might have contributed to the baseline variability in SMA-M1 modulation in the current study. Test-retest reliability of SMA-M1 ratios has previously been established for healthy younger and older adults, with moderate intraclass correlation coefficients (ICC) found for SMA-M1 modulation at ISI of 7 ms (Rurak et al., 2021a). To date, test-retest reliability of SMA-M1 modulation has not been examined in PD (see Supplementary Material Section S8 for an exploratory ICC analysis of the current data); it is important for future research to determine the test-retest reliability of dual-site TMS measures of SMA-M1 modulation in individuals with PD. A multi-modal approach using diffusion tensor imaging and dual-site TMS will also provide valuable insight into how structural connections between SMA and M1 contribute to SMA-M1 modulation in individuals with PD.

Given the difference in SMA-M1 ratios at baseline between the two sessions, we examined whether baseline SMA-M1 modulation might be associated with ccPAS-induced changes in SMA-M1 modulation. This exploratory analysis showed smaller baseline SMA-M1 ratios were significantly associated with larger increases in SMA-M1 ratios after real ccPAS (Figure 7). This also indicates that there were smaller ccPAS-induced increases in SMA-M1 ratios when baseline SMA-M1 ratios were higher. We speculate that this might be due to ceiling effects, such that ccPAS cannot increase SMA-M1 modulation beyond a certain level of facilitation. As ccPAS is believed to induce STDP, the current findings might indicate baseline SMA-M1 facilitation saturation prevents further potentiation from ccPAS. Future research should systematically explore whether potential SMA-M1 facilitation saturation affects induction of STDP by ccPAS. For example, setting stimulation parameters to induce low or high SMA-M1 facilitation at baseline, and observing the subsequent effects of ccPAS.

Alternatively, the significant increase in SMA-M1 ratios for real ccPAS might reflect regression to the mean, with the observed changes in ratios resulting from random fluctuations rather than real ccPAS effects (Barnett et al., 2005). Specifically, baseline ratios could have been unusually low during real ccPAS sessions, with ratios normalising at post real ccPAS. To reduce regression to the mean effects, future studies could consider using multiple baseline measurements and apply a selection criterion (a cut-off ratio value) using the mean of the measurements (Barnett et al., 2005).

### 4.2. ccPAS Did Not Modulate Tremor Severity

The current results showed no stimulation-specific changes in tremor severity and no significant correlations between the change in SMA-M1 ratios following ccPAS and the change in tremor severity following ccPAS. This is somewhat surprising given the significant increase in SMA-M1 modulation following ccPAS in the current study. It is possible that SMA-M1 modulation alone does not underpin resting tremor severity, at least not when participants are OFF levodopa. Indeed, baseline SMA-M1 ratios were not significantly associated with baseline resting tremor either during real or sham ccPAS sessions in the current study. This is in line with previous findings of no significant correlation between SMA-M1 modulation and tremor power for PD participants when they were OFF levodopa (Rurak et al., 2024). Previous research has shown that stronger SMA-M1 facilitation is significantly associated with lower tremor severity in individuals with PD when participants were ON levodopa (Rurak et al., 2024) and stronger blood-oxygen-level-dependent connectivity within the sensorimotor network when participants were ON levodopa compared to OFF levodopa (Caspers et al., 2021). Together, these previous findings suggest that levodopa has a normalising effect on sensorimotor network activity (Caspers et al., 2021). It is possible that ccPAS-alone cannot sufficiently modulate SMA-M1 facilitation to change tremor severity. The current study tested participants in the OFF state to examine ccPAS effects independent of levodopa, which could inform alternative interventions with fewer side effects than levodopa. It is possible that SMA-M1 ccPAS might be more efficacious if paired with levodopa. Future studies should therefore investigate the effects of ccPAS when participants are ON vs OFF levodopa.

Finally, a single session of ccPAS might be insufficient to elicit significant changes in tremor severity. In a recent study, a multi-dose ccPAS protocol (three ccPAS sessions delivered 50 minutes apart) to M1 and PPC led to significantly greater increases in cortical excitability (across single– and dual-site trials) compared with single-dose ccPAS (Goldenkoff et al., 2024). It is thus possible that multiple sessions of SMA-M1 ccPAS could elicit greater improvements in SMA-M1 modulation and lead to significant changes in resting tremor severity. The efficacy and safety of multiple ccPAS sessions has not been well-investigated (Hernandez-Pavon et al., 2023), and recommendations for TMS-based treatments for PD motor symptoms have been developed only for repetitive TMS (Lefaucheur et al., 2020). Future research should thus explore the efficacy and safety of multi-session ccPAS for reducing PD motor symptoms.

There was a trend towards an increase in tremor severity after both real and sham ccPAS. Each experimental session lasted around 2 hours; the trend towards increased tremor could be due to fatigue, which has been shown to worsen PD motor symptoms (Zhou et al., 2023). Increasing time OFF levodopa might also have contributed to the trend towards increased tremor (Chou et al., 2018). Importantly, the increase in tremor occurred for both ccPAS conditions, thus indicating that there were no specific effects of real ccPAS on tremor severity.

### 4.3. Limitations

As we were able to recruit only 14 participants (with *n*=10 for SMA-M1 data), the current study potentially is underpowered. A priori power analysis was not performed to inform our study as power analysis for linear mixed models (which was used to analyse SMA-M1 ratios in our study) requires pilot data or the specification of fixed and random parameters based on existing literature (P. Green & MacLeod, 2016), neither of which was available. Based on the effect size in the current study, a sample size of 23 participants would be required to detect significant STIMULATION X TIME interactions at 80% power. This estimated sample size could be used in future studies to verify the effects of ccPAS on SMA-M1 activity.

The baseline difference in SMA-M1 ratios between the real and sham ccPAS conditions represents a potential confound. Future research should therefore assess the test-retest reliability of TMS measures of SMA-M1 modulation in PD and examine how baseline ratios influence ccPAS effects on SMA-M1 faciliation. The significant increase in background EMG activity (pre-TMS RMS) during real ccPAS sessions should also be considered when interpreting the current findings. Although background EMG activity was not significantly associated with MEP amplitude before sham ccPAS, nor after real or sham ccPAS (see Supplementary Material Section S4), it may still represent a confound in the data. It is unclear why background EMG activity increased only during real ccPAS sessions, but this was not accompanied by changes in tremor severity and post-stimulation background EMG activity was similar after real and sham ccPAS. However, the potential confounding effects of the increased background EMG activity, as well as those from the baseline differences in SMA-M1 ratios, might be further amplified by the study’s limited sample size (Elkins, 2015). In light of these methodological limitations, the present findings should not be interpreted as robust evidence of the effects of SMA-M1 ccPAS on SMA-M1 facilitation and tremor severity, but rather as preliminary data informing the development of future hypotheses.

Individual MRIs were not used for neuronavigation and SMA localisation. The target site for SMA was located by measuring 4 cm anterior to the vertex (Cz+4), which was previously verified using individual brain anatomy (Arai et al., 2011, 2012). However, we acknowledge that the use of individual MRIs would be ideal due to potential interindividual anatomical differences. Indeed, a recent study found that Cz+4 was potentially situated over pre-supplementary motor area (pre-SMA) instead of SMA for a small number of participants (Tan et al., 2025). It is thus possible that the lack of significant changes to tremor severity was because the pre-SMA was stimulated instead of SMA for a few participants in the current study.

Finally, neuronavigation was used to monitor coil position during TMS measures of SMA-M1 modulation, but not during ccPAS. Therefore, the accuracy of coil positioning over M1 and the SMA during cPAS could not be verified and potential inaccuracies in coil placement during stimulation might have influenced the current results. Future research using neuronavigation during ccPAS delivery is needed to confirm and better characterize the effects of SMA-M1 ccPAS.

### 4.4. Conclusion

We found preliminary evidence that SMA-M1 ccPAS can increase SMA-M1 facilitation in tremor-dominant PD. However, there was no ccPAS-induced change in tremor severity, and SMA-M1 modulation was not associated with tremor severity. Future studies should explore the test-retest reliability of SMA-M1 modulation in individuals with PD ON and OFF levodopa, and whether intra-individual variability in baseline SMA-M1 facilitation impacts ccPAS effects. It is also important to note that the significant changes in SMA-M1 ratios might be due to regression to the mean; future research is needed to replicate the current findings using multiple baseline measurements. Future studies should also investigate the effects of ccPAS when participants are ON levodopa, and whether multiple sessions would be more efficacious than single sessions in alleviating resting tremor.

## Author Contribution

**JT:** Conceptualization, Methodology, Formal analysis, Investigation, Software, Data Curation, Writing – Original Draft. **BKR:** Conceptualization, Software, Writing – Review & Editing. **RCH:** Conceptualization, Methodology, Writing – Review & Editing. **JPR:** Conceptualization, Writing – Review & Editing. **BDP**: Conceptualization, Writing – Review & Editing. **PD**: Conceptualization, Writing – Review & Editing. **HF**: Conceptualization, Writing – Review & Editing. **AMV:** Conceptualization, Methodology, Formal analysis, Writing – Review & Editing, Supervision.

## Data Availability

All data produced in the present study are available upon reasonable request to the authors

## Abbreviations

PD: Parkinson’s disease
ccPAS: cortico-cortical paired associative stimulation
SMA: supplementary motor area
M1: primary motor cortex
CS: conditioning stimulus
ISI: interstimulus interval
TS: test stimulus
MEP: motor evoked potential
STDP: spike-timing-dependent plasticity
LTP: long-term potentiation
LTD: long-term depression
FDI: first dorsal interosseous
ECR: extensor carpi radialis
FCR: flexor carpi radialis

## Supplementary Material

### S1. Participant characteristics

Table S1 shows the demographic and clinical characteristics of the participants in the current study. Age and disease duration are presented in range to limit the identification of participant.

**Table S1.** Demographic and clinical characteristics of participants.

| Participant | Age (years)<sup>#</sup> | Sex | Disease duration in years<sup>#</sup> | Tremor dominant upper limb | Levodopa medication (mg) | Levodopa dose per day | Levodopa equivalent daily dose<sup>1</sup> (mg) | Other medication | MDS-UPDRS part III<sup>2</sup> |
| --- | --- | --- | --- | --- | --- | --- | --- | --- | --- |
| 1 | 70-75 | M | 5-10 | Left | 200 | 3 | 600 | Pramipexole 0.25 mg<br>Allopurinol alphapharm<br>Candesartan cilxetil<br>Atorvastatin<br>Tamsulosin | 5 |
| 2 | 70-75 | F | 1-5 | Left | 100 | 2 | 200 | Pramipexole 3 mg | 3 |
| 3 | 65-70 | M | 1-5 | Right | 200 | 2 | 400 | Safinamide 100 mg<br>Pantoprazole<br>Aspirin<br>Ezetimibe/Atorvastatin | 5 |
| 4 | 55-60 | F | 1-5 | Left | 100 | 4 | 400 | Rotigotine 12 mg | 5 |
| 5 | 60-65 | M | 1-5 | Right | 250 | 3 | 750 | Nil | 4 |
| 6 | 70-75 | M | 1-5 | Left | 200 | 2.25 | 450 | Nil | 5 |
| 7 | 60-65 | F | 1-5 | Right | 150 | 3 | 450 | Nil | 2 |
| 8 | 65-70 | M | 5-10 | Right | 200 x 1<br>75 x 2 | 3 | 350 | Pramipexole 3 mg<br>Amantadine<br>Clonazepam<br>Mirtazapine | 2 |
| 9 | 65-70 | M | 1-5 | Left | 150 | 3 | 450 | Pramipexole 3 mg | 3 |
| 10 | 60-65 | F | 1-5 | Right | 100 | 4 | 400 | Opicapone<br>Escitalopram<br>Dulaglutide | 5 |
| 11 | 70-75 | M | 1-5 | Right | 150 | 3 | 450 | Dutasteride | 5 |
| 12 | 60-65 | M | 1-5 | Right | 100 | 3 | 300 | Pramipexole 1.5 mg | 4 |
| 13 | 55-60 | M | 5-10 | Right | 200 | 4 | 800 | Sertraline<br>Clonazepam,<br>Symmetrel<br>Safinamide<br>Perindopril<br>Erbumine | 6 |
| 14 | 75-80 | F | 5-10 | Left | 100 | 4 | 400 | Nil | 3 |

*Note.* F: female; M: male; <sup>#</sup>Ages and duration of disease are presented in range to limit identification of participants. <sup>1</sup>Sum of each parkinsonian medication converted into levodopa equivalent dose. <sup>2</sup>Sum of tremor scores from the MDS-UPDRS subitems for resting tremor amplitude (item 17) and constancy (item 18) in the affected upper-limb (maximum score is 24).

### S2. Resting Tremor During Provocation Task

Resting tremor during the provocation task (provocation tremor) was analysed using a linear mixed models LMM with fixed factors of STIMULATION (real, sham), TIME (pre, post), and MUSCLE (FDI, ECR, FCR).

Figure S1 shows the provocation tremor power for each stimulation condition at each timepoint. The LMM analysis of provocation tremor power found no significant interaction effects or main effects for STIMULATION, TIME, and MUSCLE (*F*s ≤ 3.31, *p*s ≥ .071).

**Figure S1.**
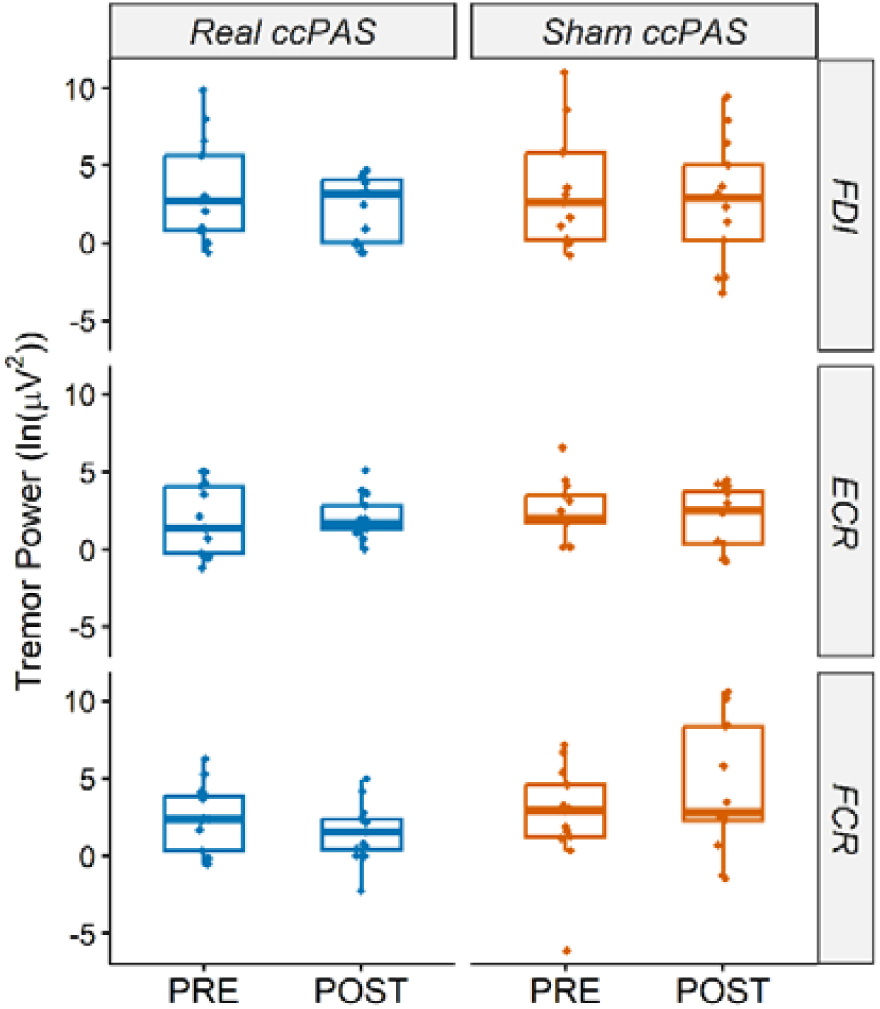
Provocation Tremor Power.

The correlation between pre-post changes in SMA-M1 ratios and pre-post changes in provocation tremor power was analysed using Spearman’s rank correlation coefficient (ρ). The correlation between baseline SMA-M1 ratios and baseline provocation tremor power was also analysed using Spearman’s ρ. The correlation analyses were conducted separately for each muscle and stimulation type.

There was no significant correlation between pre-post changes in SMA-M1 ratios and pre-post changes in provocation tremor power for both real and sham ccPAS sessions (see Figure S2). The correlation between baseline SMA-M1 ratios and baseline provocation tremor power was also not significant for both real and sham ccPAS sessions (Figure S3).

**Figure S2.**
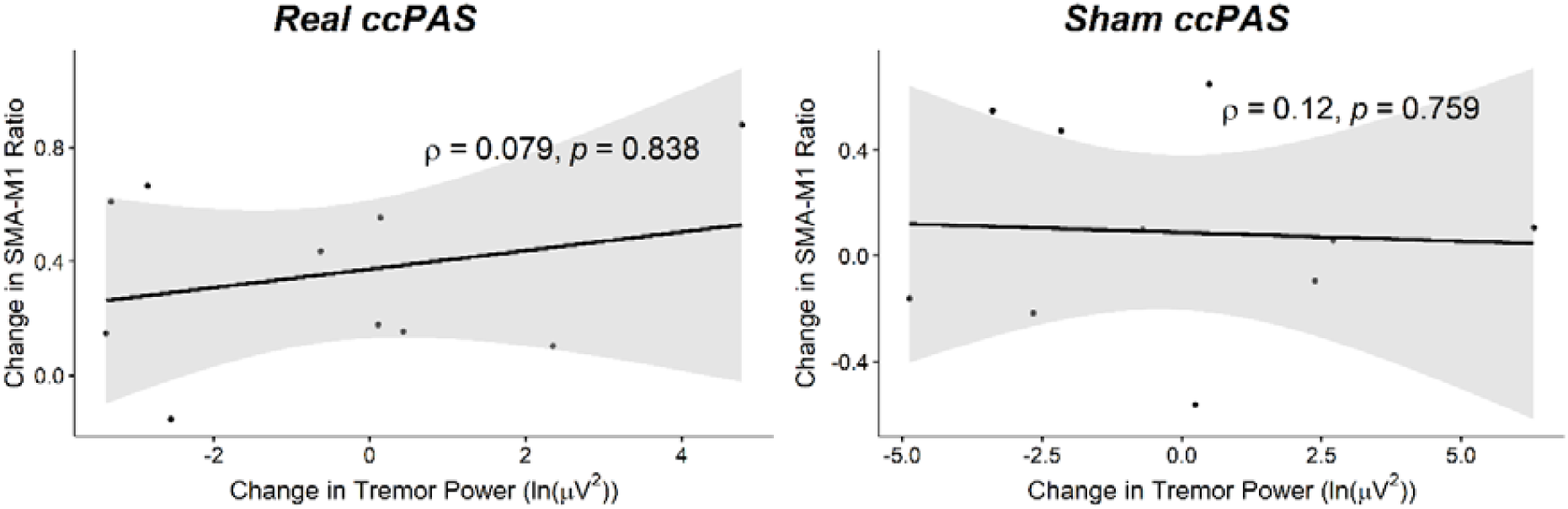
Scatterplots for Correlation between Pre-Post Changes in SMA-M1 Ratios and Pre-Post Changes in Provocation Tremor Power.

**Figure S3.**
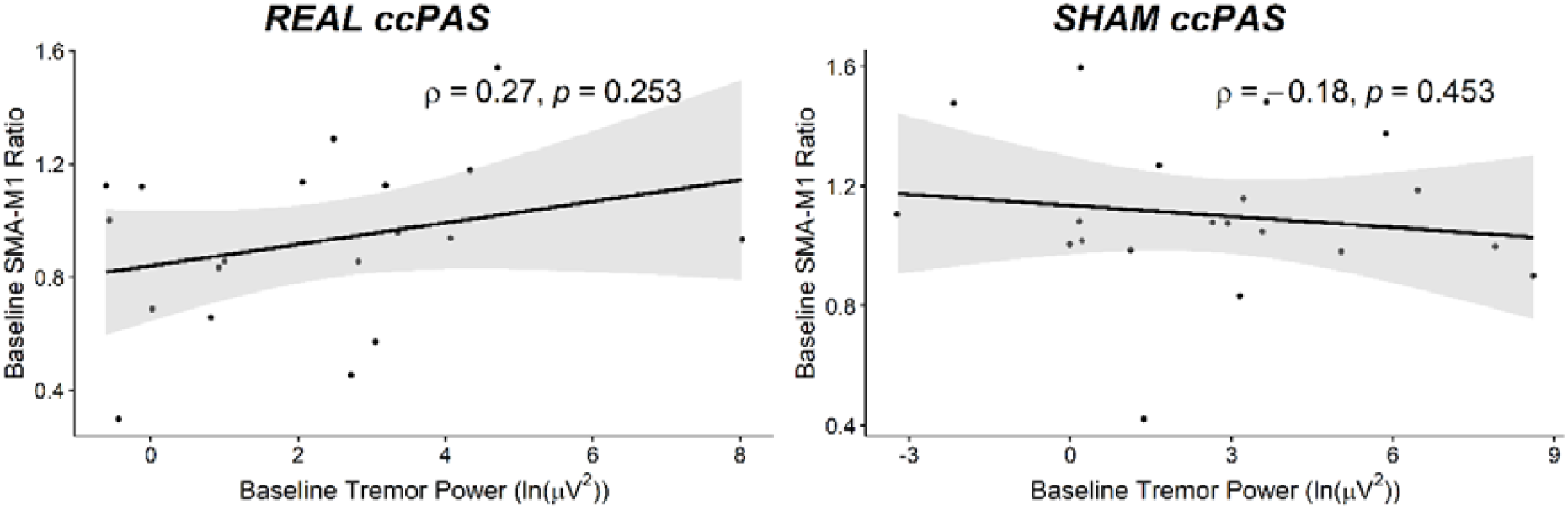
Scatterplots for Correlation between Baseline SMA-M1 Ratios and Baseline Provocation Tremor Power.

### S3. SMA-M1 Ratios: ECR and FCR Muscles

SMA-M1 ratios derived from the EMG activity measured from ECR and FCR muscles were analysed using a LMM with fixed factors of STIMULATION (real, sham) and TIME (pre, post).

### S3.1. ECR SMA-M1 Ratios

Figure S4 shows the SMA-M1 ratios for the ECR muscle for each stimulation condition at each timepoint. The LMM analysis of SMA-M1 ratios found a significant STIMULATION X TIME interaction (*F*(1, 11.11) = 7.20, *p* = .021). Post-hoc analyses showed no significant changes in SMA-M1 ratios after real ccPAS (*t* = –1.43, *p* = .169, *d* = –0.77), nor sham ccPAS (*t* = 1.89, *p* = .073, *d* = 1.02). Post-hoc contrasts also found significantly lower baseline SMA-M1 ratios for real ccPAS compared with sham ccPAS (*t* = –2.44, *p* = .024, *d* = –1.43). Post-ccPAS SMA-M1 ratios were not significantly different between real and sham ccPAS sessions (*t* = 0.61, *p* = .546, *d* = 0.36). There were no significant main effects of TIME (*F*(1, 11.11) = 0.09, *p* = .774), nor STIMULATION (*F*(1, 11.11) = 1.24, *p* = .290).

**Figure S4.**
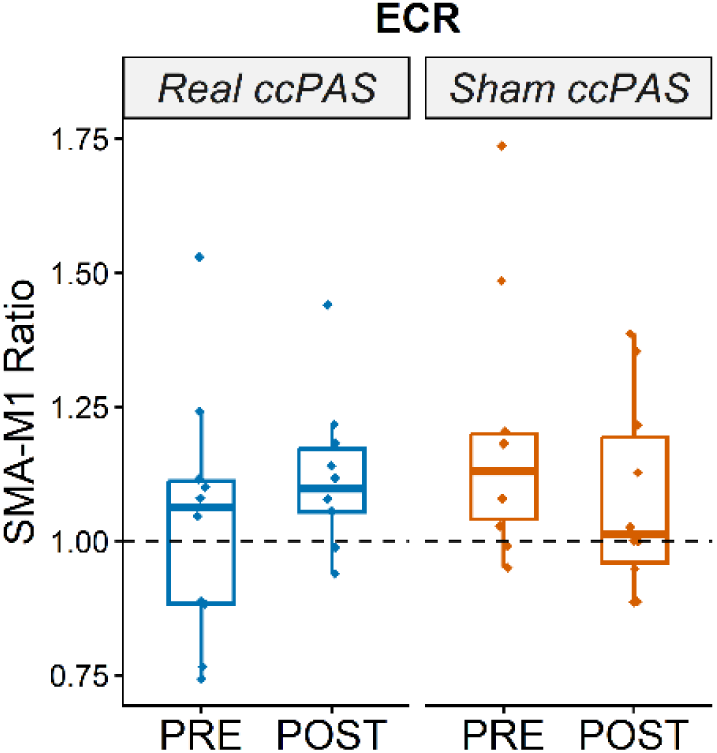
ECR SMA-M1 Ratios.

### S3.2. FCR SMA-M1 Ratios

Figure S5 shows the SMA-M1 ratios for the FCR muscle for each stimulation condition at each timepoint. The LMM analysis of SMA-M1 ratios found no significant STIMULATION X TIME interaction (*F*(1, 11.11) = 0.91, *p* = .362), and no significant main effects of TIME (*F*(1, 11.11) = 0.03, *p* = .868), nor STIMULATION (*F*(1, 11.11) = 0.07, *p* = .804).

**Figure S5.**
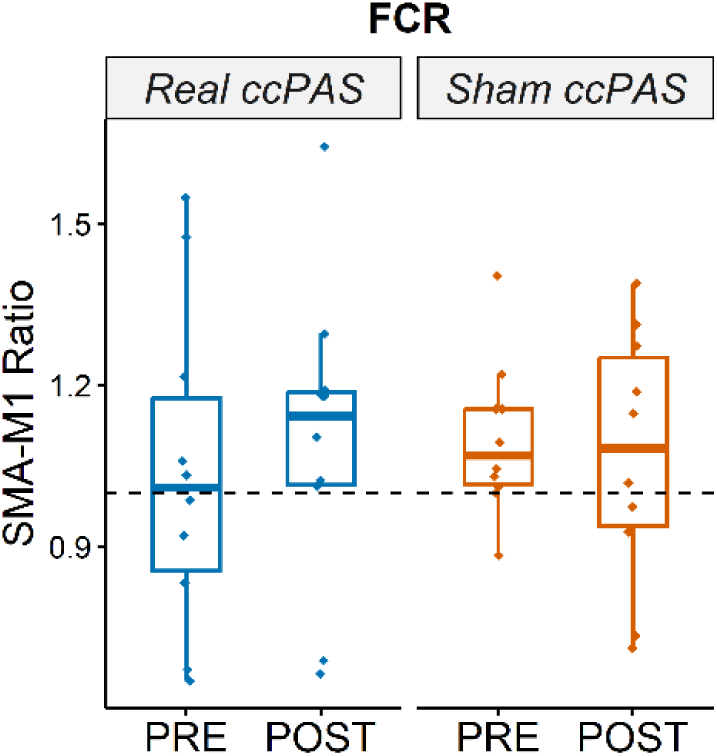
FCR SMA-M1 Ratios.

### S4. Correlation Between pre-TMS RMS and MEP Amplitudes

As there was a significant increase in pre-TMS RMS during real ccPAS sessions, we also analysed the correlation between pre-TMS RMS and MEP amplitudes using Spearman’s ρ for each trialtype (SI_1mV_-alone and dual-site trials) and stimulation condition (real and sham ccPAS). During real ccPAS sessions (see Figure S6), larger pre-TMS RMS was significantly associated with larger MEP amplitudes for SI_1mV_-alone (ρ = 0.20, *p* < .001) and dual-site trials (ρ = 0.19, *p* = .001.) at baseline but not after real ccPAS (ρ ≤ 0.058, *p*s ≥ .329). Pre-TMS RMS was not associated with MEP amplitudes before or after sham ccPAS for SI_1mV_-alone and dual-site trials (ρ ≤ 0.11, *p*s ≥ .084; see Figure S7).

**Figure S6.**
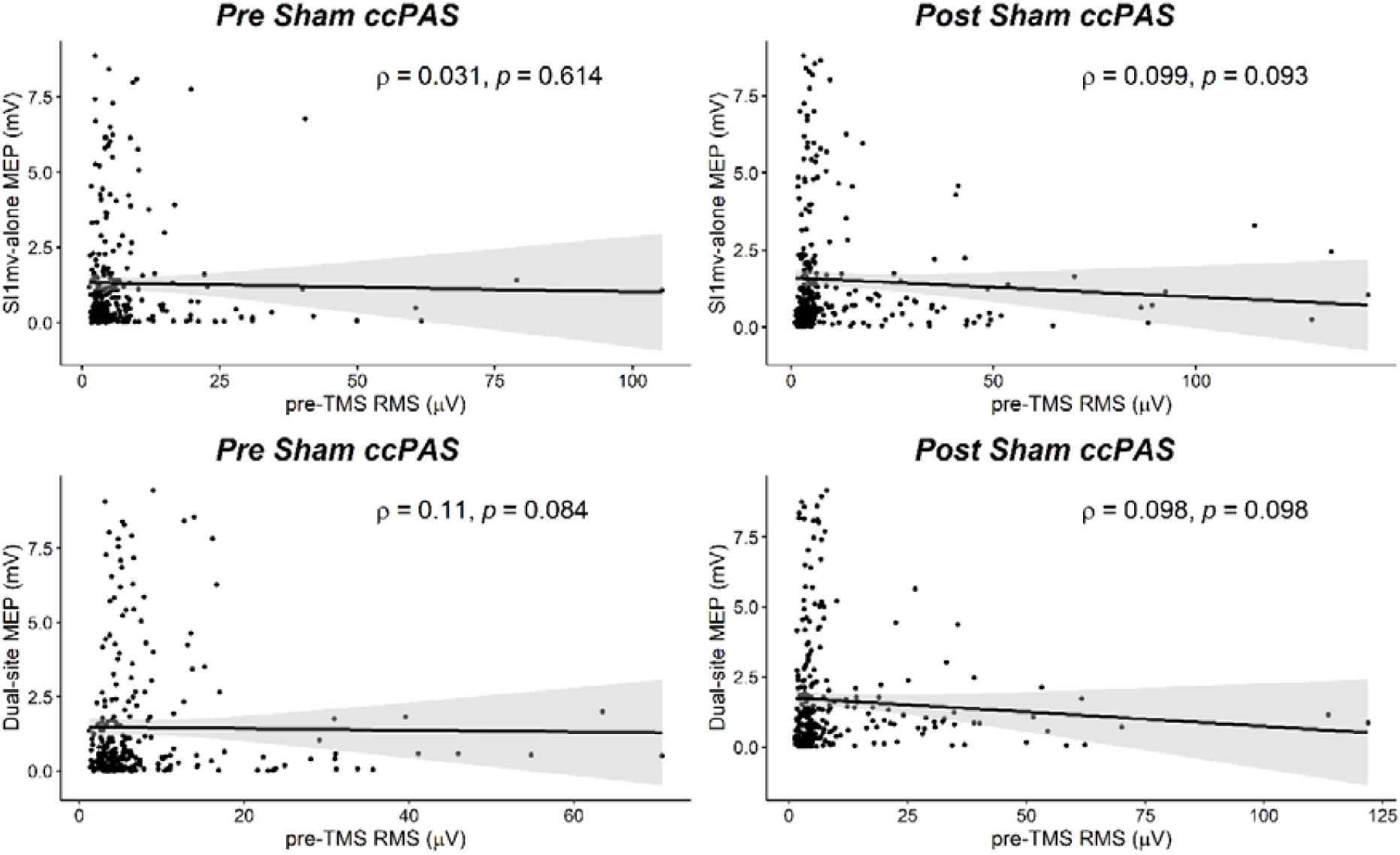
Scatterplots for Correlation between Pre-TMS RMS and MEP Amplitudes for Real ccPAS Sessions. *Note*. Larger pre-TMS RMS was significantly associated with larger MEP amplitudes for SI_1mV_-alone and dual-site trials before real ccPAS (graphs on the left). There were no signficiant correlations between pre-TMS RMS and MEP amplitudes for SI_1mV_-alone and dual-site trials after real ccPAS (graphs on the right).

**Figure S7.**
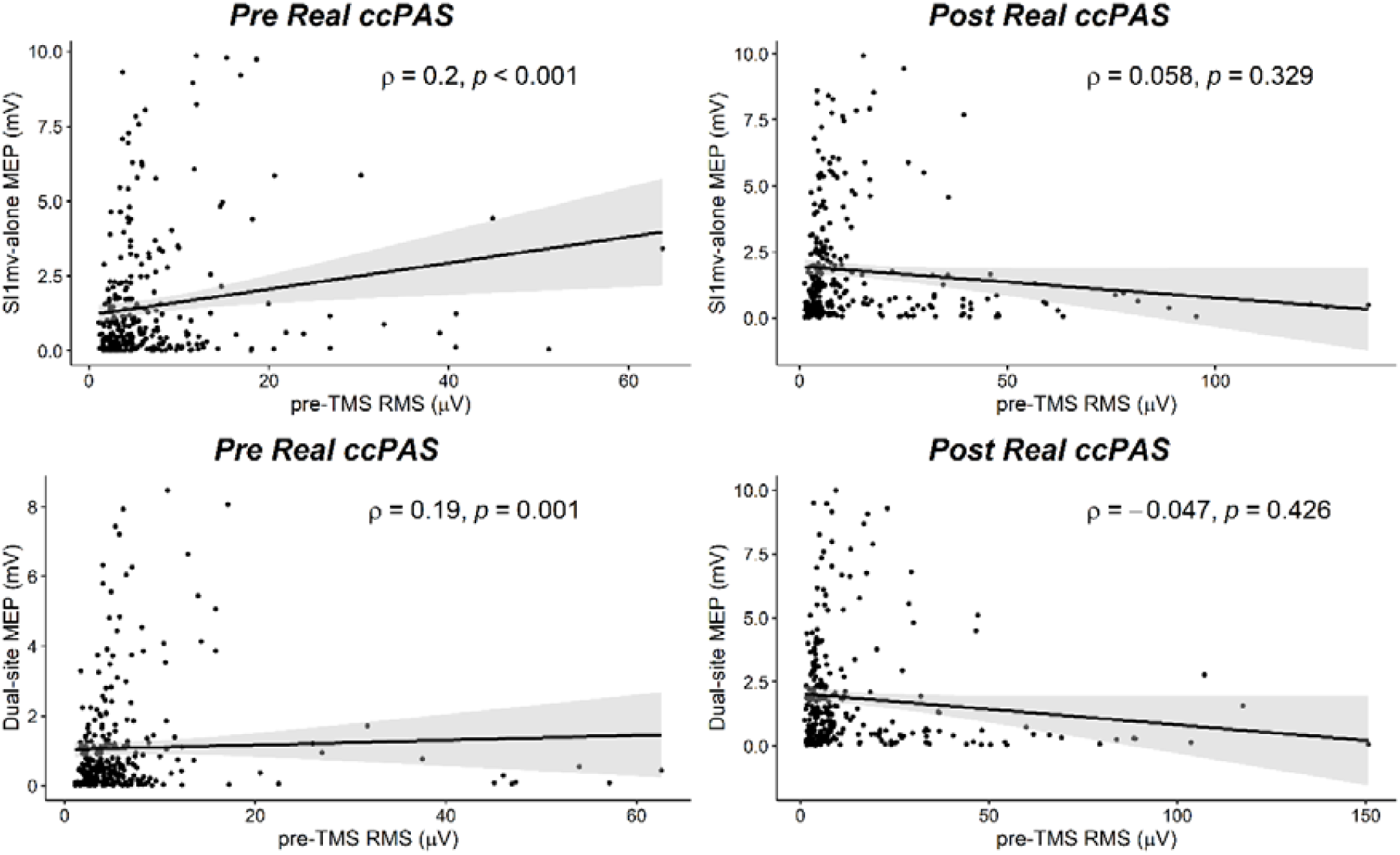
Scatterplots for Correlation between Pre-TMS RMS and MEP Amplitudes for Sham ccPAS Sessions. *Note*. There were no signficiant correlations between pre-TMS RMS and MEP amplitudes for SI_1mV_-alone and dual-site trials before or after sham ccPAS.

### S5. Resting Tremor – 10 Participants

SMA-M1 ratios were analysed for 10 participants; data from four participants could not be analysed due to a technical error. A LMM analysis was conducted to analyse the resting tremor power for these 10 participants whose SMA-M1 ratios could be analysed, with fixed factors of STIMULATION (real, sham), TIME (pre, post), and MUSCLE (FDI, ECR, FCR). The analysis found a significant main effect of TIME (F(1, 111.11) = 4.72, p = .032), with a pre-post increase in resting tremor power across both real and sham ccPAS sessions. There were no other significant interaction effects or main effects (Fs ≤ 2.93, ps ≥ .056). Figure S8 shows the resting tremor power for the 10 participants at each muscle, before and after real and sham ccPAS.

**Figure S8.**
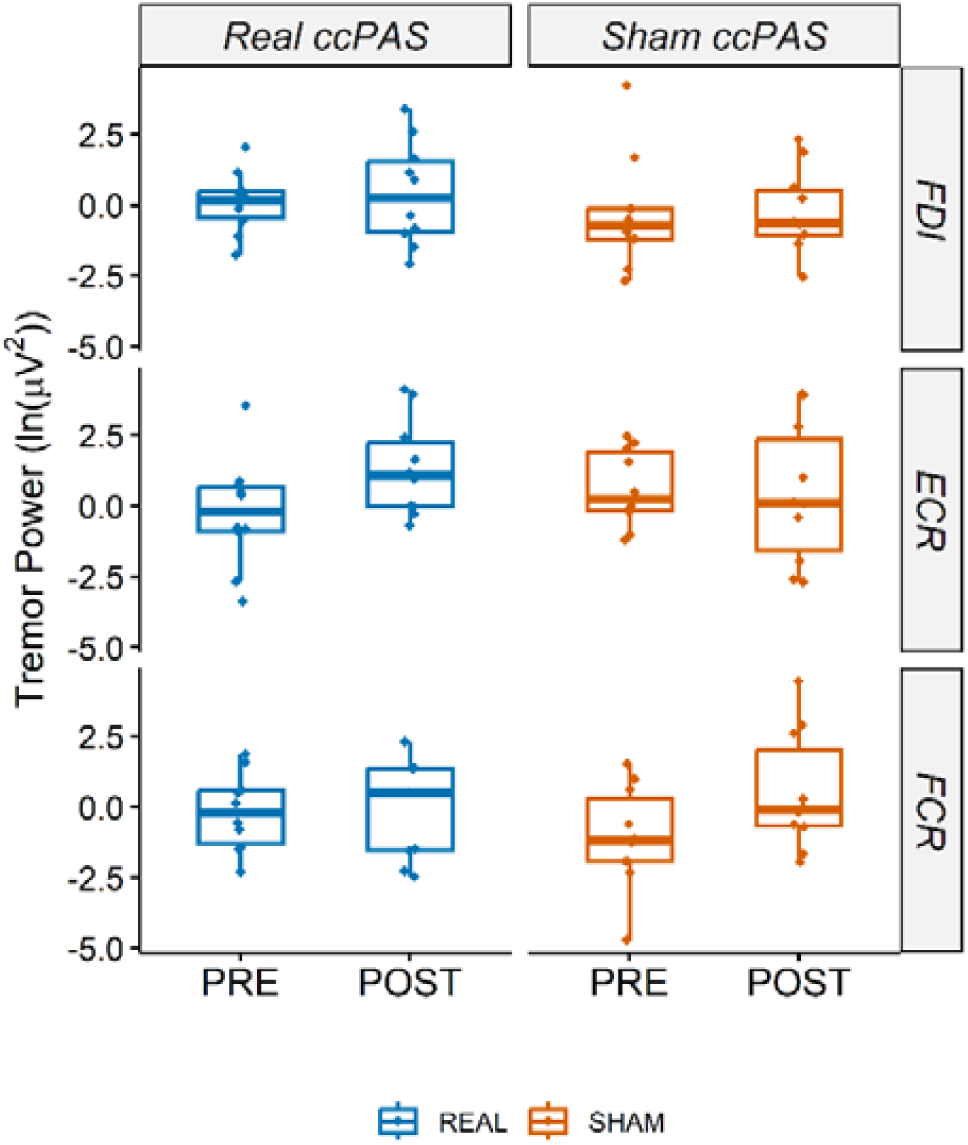
Resting Tremor Power for 10 Participants. *Note*. There were no stimulation-specific changes in resting tremor power for the 10 participants whose SMA-M1 ratios were analysed.

### S6. Baysesian Linear Mixed Model Analysis of Resting Tremor

We performed a supplementary analysis of tremor power using a Bayesian linear mixed model with fixed factors of STIMULATION (real, sham), TIME (pre, post), and MUSCLE (FDI, ECR, FCR) with both random intercepts and slopes. A weakly informative prior was specified for the fixed effects with a Gaussian distribution mean of 0 and standard deviation of 1 (N(0,1)). A posterior distribution over possible parameter values was sampled using Markov chain Monte Carlo (MCMC) sampling NUTS (No-U-Turn Sampler) algorithm. We ran 4 separate chains with 6000 iterations each, with the initial 1000 iterations discarded as warm-ups, and with thinning set to select every 5^th^ sample in the chain.

The results indicated a good convergence of the model, with all R-hat values = 1.00, and adequate effective sample sizes (ESS; Bulk ESS ≥ 3635; Tail ESS ≥ 3603). However, the model revealed inconclusive effects for 3-way interactions (pds ≤ 82.1%; all HDI in ROPE between 8.24 and 9.84%), and thus the null hypothesis was not accepted or rejected.

Similarly, the STIMULATION x TIME interaction was inconclusive (pd = 74.8%; 12.24% HDI in ROPE). These findings indicate that there is no clear evidence for or against a practically meaningful effect of ccPAS on tremor power across stimulation types and muscles.

### S7. Correlation Between SMA-M1 Ratio and Tremor Severity

There was no significant correlation between pre-post changes in SMA-M1 ratios and pre-post changes in tremor power for both real and sham ccPAS sessions (ρ ≤ 0.13, *p*s ≥ .733; see Figure S9). The correlation between baseline SMA-M1 ratios and baseline tremor power was also not significant for both real and sham ccPAS sessions (ρ ≤ –0.39, *p*s ≥ .263; Figure S10).

**Figure S9.**
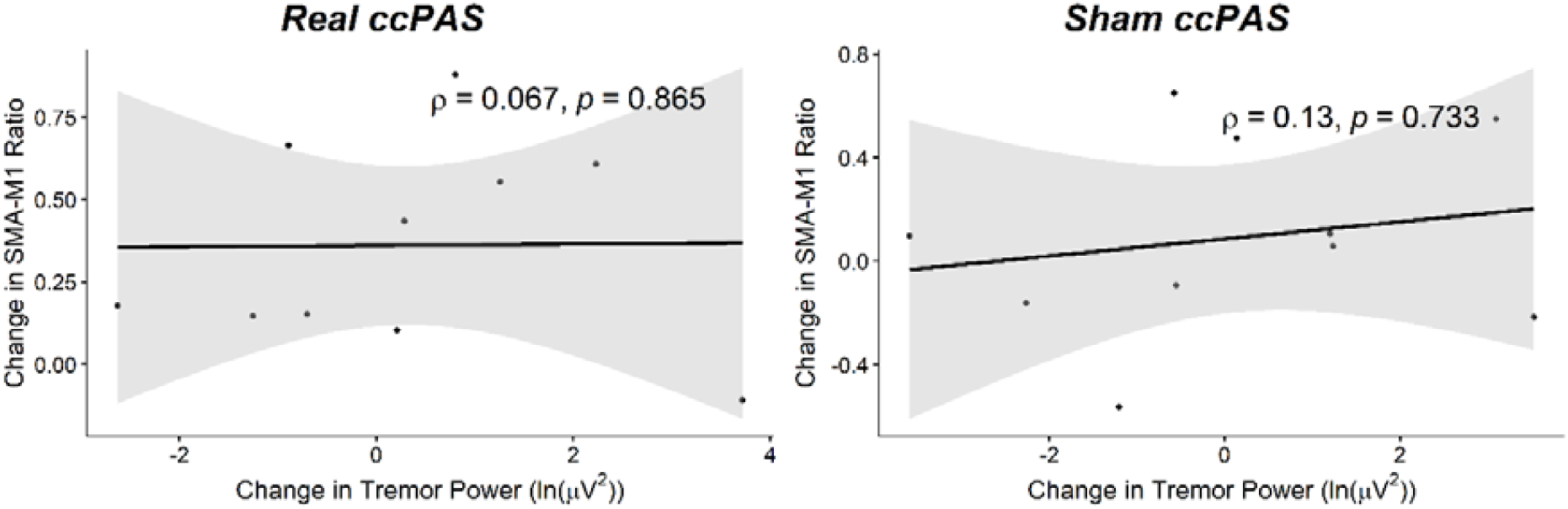
Scatterplots for Correlation between Pre-Post Changes in SMA-M1 Ratios and Pre-Post Changes in Tremor Power. *Note*. There was no significant correlation between pre-post changes in SMA-M1 ratios and pre-post changes in tremor power for both real and sham ccPAS sessions.

**Figure S10.**
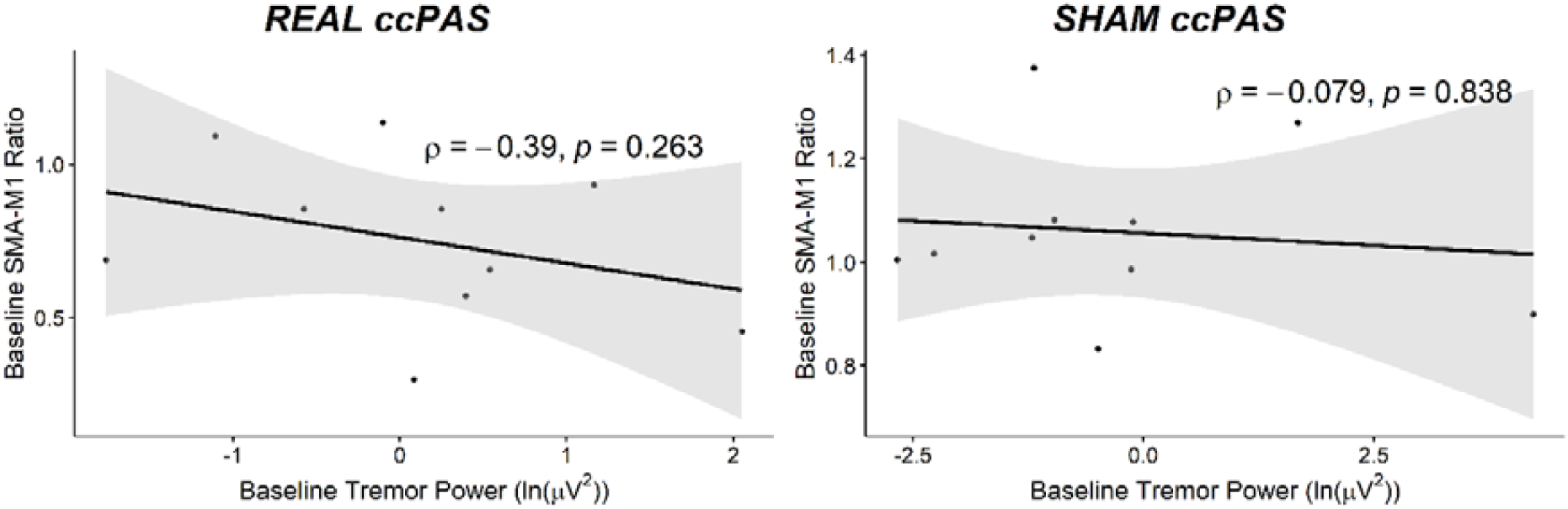
Scatterplots for Correlation between Baseline SMA-M1 Ratios and Baseline Tremor Power. *Note*. Baseline SMA-M1 ratios were not significantly associated with baseline tremor power for both real and sham ccPAS sessions.

### S8. Exploratory Test-Retest Reliability Analyses

Despite the small sample size of the current study, we calculated intraclass correlation coefficients (ICC) using a single-rating, absolute agreement, two-way fixed effects model (Koo & Li, 2016) to explore the test-retest reliability of baseline SMA-M1 ratios. The results (ICC value of –0.06 (95% CI[-0.32 0.41]) indicate zero reliability (Bartko, 1976). However, we note that the current sample of 10 participants is too small to run a sufficiently powered ICC analysis; indeed, a minimum sample size of 74 participants is required for an estimated ICC value of 0.5 (computed using Mondal et al.’s (2024) R Shiny app, with the Wald confidence interval method with Fisher variance).

## References

Akkal, D., Dum, R. P., & Strick, P. L. (2007). Supplementary Motor Area and Presupplementary Motor Area: Targets of Basal Ganglia and Cerebellar Output. The Journal of Neuroscience, 27(40), 10659–10673. 10.1523/JNEUROSCI.3134-07.2007

Arai, N., Lu, M.-K., Ugawa, Y., & Ziemann, U. (2012). Effective connectivity between human supplementary motor area and primary motor cortex: A paired-coil TMS study. Experimental Brain Research, 220(1), 79–87. 10.1007/s00221-012-3117-5

Arai, N., Müller-Dahlhaus, F., Murakami, T., Bliem, B., Lu, M.-K., Ugawa, Y., & Ziemann, U. (2011). State-Dependent and Timing-Dependent Bidirectional Associative Plasticity in the Human SMA-M1 Network. Journal of Neuroscience, 31(43), 15376–15383. 10.1523/JNEUROSCI.2271-11.2011

Barnett, A. G., van der Pols, J. C., & Dobson, A. J. (2005). Regression to the mean: What it is and how to deal with it. International Journal of Epidemiology, 34(1), 215–220. 10.1093/ije/dyh299

Barr, D. J., Levy, R., Scheepers, C., & Tily, H. J. (2013). Random effects structure for confirmatory hypothesis testing: Keep it maximal. Journal of Memory and Language, 68(3), 255–278. 10.1016/j.jml.2012.11.001

Bates, D., Mächler, M., Bolker, B., & Walker, S. (2015). Fitting Linear Mixed-Effects Models Using lme4. Journal of Statistical Software, 67(1), 1–48. 10.18637/jss.v067.i01

Bates, D., Maechler, M., Bolker, B., & Walker, S. (2025). lme4: Linear Mixed-Effects Models using Eigen and S4. https://github.com/lme4/lme4/

Belyk, M., Murphy, B. K., & Beal, D. S. (2019). Accessory to dissipate heat from transcranial magnetic stimulation coils. Journal of Neuroscience Methods, 314, 28–30. 10.1016/j.jneumeth.2019.01.008

Bi, G., & Poo, M. (1998). Synaptic Modifications in Cultured Hippocampal Neurons: Dependence on Spike Timing, Synaptic Strength, and Postsynaptic Cell Type. Journal of Neuroscience, 18(24), 10464–10472. 10.1523/JNEUROSCI.18-24-10464.1998

Bloem, B. R., Okun, M. S., & Klein, C. (2021). Parkinson’s disease. The Lancet, 397(10291), 2284–2303. 10.1016/S0140-6736(21)00218-X

Boylan, L. S., Pullman, S. L., Lisanby, S. H., Spicknall, K. E., & Sackeim, H. A. (2001). Repetitive transcranial magnetic stimulation to SMA worsens complex movements in Parkinson’s disease. Clinical Neurophysiology, 112(2), 259–264. 10.1016/S1388-2457(00)00519-8

Carson, N., Leach, L., & Murphy, K. J. (2018). A re-examination of Montreal Cognitive Assessment (MoCA) cutoff scores. International Journal of Geriatric Psychiatry, 33(2), 379–388. 10.1002/gps.4756

Caspers, J., Rubbert, C., Eickhoff, S. B., Hoffstaedter, F., Südmeyer, M., Hartmann, C. J., Sigl, B., Teichert, N., Aissa, J., Turowski, B., Schnitzler, A., & Mathys, C. (2021). Within– and across-network alterations of the sensorimotor network in Parkinson’s disease. Neuroradiology, 63(12), 2073–2085. 10.1007/s00234-021-02731-w

Chou, K. L., Stacy, M., Simuni, T., Miyasaki, J., Oertel, W. H., Sethi, K., Fernandez, H. H., & Stocchi, F. (2018). The spectrum of “off” in Parkinson’s disease: What have we learned over 40 years? Parkinsonism & Related Disorders, 51, 9–16. 10.1016/j.parkreldis.2018.02.001

Cohen, J. (1988). Statistical Power Analysis for the Behavioral Sciences. Routledge. 10.4324/9780203771587

Dirkx, M. F., & Bologna, M. (2022). The pathophysiology of Parkinson’s disease tremor. Journal of the Neurological Sciences, 435. 10.1016/j.jns.2022.120196

Dirkx, M. F., Zach, H., van Nuland, A. J., Bloem, B. R., Toni, I., & Helmich, R. C. (2020). Cognitive load amplifies Parkinson’s tremor through excitatory network influences onto the thalamus. Brain, 143(5), 1498–1511. 10.1093/brain/awaa083

Dum, R. P., & Strick, P. L. (2002). Motor areas in the frontal lobe of the primate. Physiology & Behavior, 77(4–5), 677–682. 10.1016/s0031-9384(02)00929-0

Elkins, M. R. (2015). Assessing baseline comparability in randomised trials. Journal of Physiotherapy, 61(4), 228–230. 10.1016/j.jphys.2015.07.005

Fitzgerald, P. B., Fountain, S., & Daskalakis, Z. J. (2006). A comprehensive review of the effects of rTMS on motor cortical excitability and inhibition. Clinical Neurophysiology, 117(12), 2584–2596. 10.1016/j.clinph.2006.06.712

Fox, J., & Weisberg, S. (2019). An R Companion to Applied Regression (Third). Sage. https://www.john-fox.ca/Companion/

Fox, J., Weisberg, S., & Price, B. (2024). car: Companion to Applied Regression. https://r-forge.r-project.org/projects/car/

Gamer, M., Lemon, J., & Singh, I. F. P. (2019). irr: Various Coefficients of Interrater Reliability and Agreement. https://www.r-project.org

Goldenkoff, E. R., Deluisi, J. A., Lee, T. G., Hampstead, B. M., Taylor, S. F., Polk, T. A., & Vesia, M. (2024). Repeated spaced cortical paired associative stimulation promotes additive plasticity in the human parietal-motor circuit. Clinical Neurophysiology, 166, 202–210. 10.1016/j.clinph.2024.08.005

Green, P. E., Ridding, M. C., Hill, K. D., Semmler, J. G., Drummond, P. D., & Vallence, A.-M. (2018). Supplementary motor area—Primary motor cortex facilitation in younger but not older adults. Neurobiology of Aging, 64, 85–91. 10.1016/j.neurobiolaging.2017.12.016

Green, P., & MacLeod, C. J. (2016). SIMR: An R package for power analysis of generalized linear mixed models by simulation. Methods in Ecology and Evolution, 7(4), 493–498. 10.1111/2041-210X.12504

Groppa, S., Oliviero, A., Eisen, A., Quartarone, A., Cohen, L. G., Mall, V., Kaelin-Lang, A., Mima, T., Rossi, S., Thickbroom, G. W., Rossini, P. M., Ziemann, U., Valls-Solé, J., & Siebner, H. R. (2012). A practical guide to diagnostic transcranial magnetic stimulation: Report of an IFCN committee. Clinical Neurophysiology, 123(5), 858–882. 10.1016/j.clinph.2012.01.010

Helmich, R. C., Van den Berg, K. R. E., Panyakaew, P., Cho, H. J., Osterholt, T., McGurrin, P., Shamim, E. A., Popa, T., Haubenberger, D., & Hallett, M. (2021). Cerebello-Cortical Control of Tremor Rhythm and Amplitude in Parkinson’s Disease. Movement Disorders, 36(7), 1727–1729. 10.1002/mds.28603

Hernandez-Pavon, J. C., San Agustín, A., Wang, M. C., Veniero, D., & Pons, J. L. (2023). Can we manipulate brain connectivity? A systematic review of cortico-cortical paired associative stimulation effects. Clinical Neurophysiology, 154, 169–193. 10.1016/j.clinph.2023.06.016

Heusinkveld, L. E., Hacker, M. L., Turchan, M., Davis, T. L., & Charles, D. (2018). Impact of Tremor on Patients With Early Stage Parkinson’s Disease. Frontiers in Neurology, 9. 10.3389/fneur.2018.00628

Jitkritsadakul, O., Jagota, P., & Bhidayasiri, R. (2016). Pathophysiology of parkinsonian tremor: A focused narrative review. Asian Biomedicine, 10(s1), s15–s22.

Kassambara, A. (2021). rstatix: Pipe-Friendly Framework for Basic Statistical Tests. https://rpkgs.datanovia.com/rstatix/

Kassambara, A. (2023). ggpubr: Ggplot2 Based Publication Ready Plots. https://rpkgs.datanovia.com/ggpubr/

Knudson, C. (2024). glmm: Generalized Linear Mixed Models via Monte Carlo Likelihood Approximation. https://CRAN.R-project.org/package=glmm

Kuznetsova, A., Brockhoff, P. B., & Christensen, R. H. B. (2017). lmerTest Package: Tests in Linear Mixed Effects Models. Journal of Statistical Software, 82(13), 1–26. 10.18637/jss.v082.i13

Lefaucheur, J.-P., Aleman, A., Baeken, C., Benninger, D. H., Brunelin, J., Di Lazzaro, V., Filipović, S. R., Grefkes, C., Hasan, A., Hummel, F. C., Jääskeläinen, S. K., Langguth, B., Leocani, L., Londero, A., Nardone, R., Nguyen, J.-P., Nyffeler, T., Oliveira-Maia, A. J., Oliviero, A., … Ziemann, U. (2020). Evidence-based guidelines on the therapeutic use of repetitive transcranial magnetic stimulation (rTMS): An update (2014–2018). Clinical Neurophysiology, 131(2), 474–528. 10.1016/j.clinph.2019.11.002

Lenth, R. V. (2023). emmeans: Estimated Marginal Means, aka Least-Squares Means. https://github.com/rvlenth/emmeans

López-Alonso, V., Cheeran, B., Río-Rodríguez, D., & Fernández-del-Olmo, M. (2014). Inter-individual Variability in Response to Non-invasive Brain Stimulation Paradigms. *Brain Stimulation: Basic*, Translational, and Clinical Research in Neuromodulation, 7(3), 372–380. 10.1016/j.brs.2014.02.004

Lu, M.-K., Chiou, S.-M., Ziemann, U., Huang, H.-C., Yang, Y.-W., & Tsai, C.-H. (2015). Resetting tremor by single and paired transcranial magnetic stimulation in Parkinson’s disease and essential tremor. Clinical Neurophysiology, 126(12), 2330–2336. 10.1016/j.clinph.2015.02.010

Luppino, G., Matelli, M., Camarda, R., & Rizzolatti, G. (1993). Corticocortical connections of area F3 (SMA-proper) and area F6 (pre-SMA) in the macaque monkey. Journal of Comparative Neurology, 338(1), 114–140. 10.1002/cne.903380109

Magee, J. C., & Johnston, D. (1997). A Synaptically Controlled, Associative Signal for Hebbian Plasticity in Hippocampal Neurons. Science, 275(5297), 209–213. 10.1126/science.275.5297.209

Mangiafico, S. (2024). rcompanion: Functions to Support Extension Education Program Evaluation. https://CRAN.R-project.org/package=rcompanion

Markram, H., Lübke, J., Frotscher, M., & Sakmann, B. (1997). Regulation of Synaptic Efficacy by Coincidence of Postsynaptic APs and EPSPs. Science, 275(5297), 213–215. 10.1126/science.275.5297.213

Muakkassa, K. F., & Strick, P. L. (1979). Frontal lobe inputs to primate motor cortex: Evidence for four somatotopically organized ‘premotor’ areas. Brain Research, 177(1), 176–182. 10.1016/0006-8993(79)90928-4

Pellegrini, M., Zoghi, M., & Jaberzadeh, S. (2020). A Checklist to Reduce Response Variability in Studies Using Transcranial Magnetic Stimulation for Assessment of Corticospinal Excitability: A Systematic Review of the Literature. Brain Connectivity, 10(2), 53–71. 10.1089/brain.2019.0715

Pirker, W., Katzenschlager, R., Hallett, M., & Poewe, W. (2023). Pharmacological Treatment of Tremor in Parkinson’s Disease Revisited. Journal of Parkinson’s Disease, 13(2), 127. 10.3233/JPD-225060

Puri, R., & Hinder, M. R. (2022). Response bias reveals the role of interhemispheric inhibitory networks in movement preparation and execution. Neuropsychologia, 165, 108120. 10.1016/j.neuropsychologia.2021.108120

Rizzo, V., Siebner, H. S., Morgante, F., Mastroeni, C., Girlanda, P., & Quartarone, A. (2009). Paired Associative Stimulation of Left and Right Human Motor Cortex Shapes Interhemispheric Motor Inhibition based on a Hebbian Mechanism. Cerebral Cortex, 19(4), 907–915. 10.1093/cercor/bhn144

Rurak, B. K., Rodrigues, J. P., Power, B. D., Drummond, P. D., & Vallence, A. M. (2021a). Test Re-test Reliability of Dual-site TMS Measures of SMA-M1 Connectivity Differs Across Inter-stimulus Intervals in Younger and Older Adults. Neuroscience, 472, 11–24. 10.1016/j.neuroscience.2021.07.023

Rurak, B. K., Rodrigues, J. P., Power, B. D., Drummond, P. D., & Vallence, A.-M. (2021b). Reduced SMA-M1 connectivity in older than younger adults measured using dual-site TMS. European Journal of Neuroscience, 54(7), 6533–6552. 10.1111/ejn.15438

Rurak, B. K., Tan, J., Rodrigues, J. P., Power, B. D., Drummond, P. D., & Vallence, A. M. (2024). Cortico-cortical connectivity is influenced by levodopa in tremor-dominant Parkinson’s disease. Neurobiology of Disease, 196, 106518. 10.1016/j.nbd.2024.106518

Sale, M. V., Ridding, M. C., & Nordstrom, M. A. (2008). Cortisol inhibits neuroplasticity induction in human motor cortex. The Journal of Neuroscience: The Official Journal of the Society for Neuroscience, 28(33), 8285–8293. 10.1523/JNEUROSCI.1963-08.2008

Schulz, R., Braass, H., Liuzzi, G., Hoerniss, V., Lechner, P., Gerloff, C., & Hummel, F. C. (2015). White matter integrity of premotor–motor connections is associated with motor output in chronic stroke patients. NeuroImage: Clinical, 7, 82–86. 10.1016/j.nicl.2014.11.006

Shima, K., & Tanji, J. (1998). Involvement of NMDA and non-NMDA receptors in the neuronal responses of the primary motor cortex to input from the supplementary motor area and somatosensory cortex: Studies of task-performing monkeys. The Japanese Journal of Physiology, 48(4), 275–290. 10.2170/jjphysiol.48.275

Singmann, H., & Kellen, D. (2019). An Introduction to Mixed Models for Experimental Psychology. In D. Spieler & E. Schumacher (Eds.), New Methods in Cognitive Psychology (pp. 4–31). Routledge. 10.4324/9780429318405

Strick, P. L. (1985). How do the basal ganglia and cerebellum gain access to the cortical motor areas? Behavioural Brain Research, 18(2), 107–123. 10.1016/0166-4328(85)90067-1

Tan, J., Rowe, G., Puri, R., Needham, M., Marneweck, M., Radia, S., & Vallence, A.-M. (2025). Investigating the effects of age and conditioning stimulation intensity on SMA–M1 connectivity in younger, middle-aged, and older adults. European Journal of Applied Physiology. 10.1007/s00421-025-05904-0

Waring, E., Quinn, M., McNamara, A., Rubia, E. A. de la, Zhu, H., & Ellis, S. (2022). skimr: Compact and Flexible Summaries of Data. https://docs.ropensci.org/skimr/

Wickham, H. (2023). tidyverse: Easily Install and Load the Tidyverse. https://tidyverse.tidyverse.org

Wickham, H., & Bryan, J. (2023). readxl: Read Excel Files. https://readxl.tidyverse.org

Zach, H., Dirkx, M. F., Roth, D., Pasman, J. W., Bloem, B. R., & Helmich, R. C. (2020). Dopamine-responsive and dopamine-resistant resting tremor in Parkinson disease. Neurology, 95(11), e1461–e1470. 10.1212/WNL.0000000000010316

Zhou, X., Xiang, Y., Song, T., Zhao, Y., Pan, H., Xu, Q., Chen, Y., Sun, Q., Wu, X., Yan, X., Guo, J., Tang, B., Lei, L., & Liu, Z. (2023). Characteristics of fatigue in Parkinson’s disease: A longitudinal cohort study. Frontiers in Aging Neuroscience, 15, 1133705. 10.3389/fnagi.2023.1133705

Zhu, H., Huang, J., Deng, L., He, N., Cheng, L., Shu, P., Yan, F., Tong, S., Sun, J., & Ling, H. (2019). Abnormal Dynamic Functional Connectivity Associated With Subcortical Networks in Parkinson’s Disease: A Temporal Variability Perspective. Frontiers in Neuroscience, 13. 10.3389/fnins.2019.00080

## References

Bartko, J. J. (1976). On various intraclass correlation reliability coefficients. Psychological Bulletin, 83(5), 762–765. 10.1037/0033-2909.83.5.762

Koo, T. K., & Li, M. Y. (2016). A Guideline of Selecting and Reporting Intraclass Correlation Coefficients for Reliability Research. Journal of Chiropractic Medicine, 15(2), 155–163. 10.1016/j.jcm.2016.02.012

Mondal, D., Vanbelle, S., Cassese, A., & Candel, M. J. (2024). Review of sample size determination methods for the intraclass correlation coefficient in the one-way analysis of variance model. Statistical Methods in Medical Research, 33(3), 532–553. 10.1177/09622802231224657

